# Epidemic Surveillance Models for Containing the Spread of SARS-CoV-2 Variants: Taiwan Experience

**DOI:** 10.1101/2021.10.19.21265107

**Authors:** Amy Ming-Fang Yen, Tony Hsiu-Hsi Chen, Wei-Jung Chang, Ting-Yu Lin, Grace Hsiao-Hsuan Jen, Chen-Yang Hsu, Sen-Te Wang, Huong Dang, Sam Li-Sheng Chen

## Abstract

**Objectives:** Two kinds of epidemic surveillance models are presented for containing the spread of SARS-CoV-2 variants so as to avert and stamp out a community-acquired outbreak (CAO) with non-pharmaceutical interventions (NPIs), tests, and vaccination.

**Design:** The surveillance of domestic cluster infections transmitted from imported cases with one-week time lag assessed by the Poisson model and the surveillance of whether, how and when NPIs and test contained the CAO with the SEIR model.

**Settings:** Border and Community of Taiwan.

**Main Outcome Measurements:** The expected number and the upper bound of the 95% credible interval (CrI) of weekly covid-19 cases compared with the observed number for assessing the threshold of a CAO; effective reproductive number (R_t_) and the effectiveness of NPIs for containing a CAO.

**Results:** For the period of January-September 2020 when the wild type and the D614G period were prevailing, an increase in one imported case prior to one week would lead to 9.54% (95% CrI 6.44% to 12.59%) higher risk of domestic cluster infection that provides a one-week prior alert signal for more stringent NPIs and active testing locally. Accordingly, there was an absence of CAO until the Alpha VOC period of February 2021. However, given level one of NPI alert the risk of domestic cluster infections was gradually elevated to 14.14% (95% CrI 5.41% to 25.10%), leading to the Alpha VOC CAOs of six hotspots around mid-May 2021. It took two-and-half months for containing this CAO mainly with level three of NPI alert and rapid test and partially by the rolling out of vaccination. By applying the SEIR model, the R_t_ decreased from 4.0 at beginning to 0.7 on 31 July 2021 in parallel with the escalating NPIs from 30% to 90%. Containing a small outbreak of Delta VOC during this CAO period was also evaluated and demonstrated. After controlling the CAO, it again returned to imported-domestic transmission for Delta VOC from July until September 2021, giving an estimate of 10.16% (95% CrI: 7.01% to 13.59%) for the risk of several small cluster infections. However, there was an absence of CAO that resulted from the effectiveness of NPIs and tests, and the rapid expansion of vaccination.

**Conclusions:** Averting and containing CAOs of SARS-CoV-2 variants are demonstrated by two kinds of epidemic surveillance models that have been applied to Taiwan scenario. These two models can be accommodated to monitor the epidemic of forthcoming emerging SARS-CoV-2 VOCs with various circumstances of vaccine coverage, NPIs, and tests in countries worldwide.

## Introduction

As of September 2021, many countries around the world have experienced repeated surges of covid-19 epidemic alternating between lifting and operating non-pharmaceutical interventions (NPIs) in the facing of lasting covid-19 pandemic before and after the era of vaccination.^1–4^ It should be noted that such a transmission is also aggravated by the emerging severe acute respiratory syndrome coronavirus 2 (SARS-CoV-2) variants, particularly Variants of Concerns (VOCs) and Variants of Interests (VOIs) that not only increase transmissibility, leading to the rapid spread from importation, household, institution, and community, but also have a higher likelihood of escaping immune response after vaccination, resulting in vaccine breakthrough.^4–7^

It is customary to see a series of repeated surges of community-acquired outbreaks (CAOs) often resulting from the spread of the imported cases of each country while quarantine and isolation of the border control are ineffective. Large-scale community covid-19 outbreaks were noted in many countries after the first wave of imported case^2,8,9^ and the subsequent global spread through inter-continental transmission even after border control measures and travel restriction operated by many countries.^2,8–14^ It is well recognized that the transmission routes lead to large-scale CAOs often begin with small cluster infections among households and institutions due to careless quarantine and isolation of importation cases that render susceptibles become infected after close contact with them. In the absence of NPIs on population level, these infectives resulting from small cluster infections may be further propagated into a large epidemic episode in the neighborhood of community and were further spread into hotspot over hotspot.^9,13,15^ This can be also observed in recent cluster infections among households and institutions aftermath of vaccine breakthrough.^6,16–18^

Once CAOs occur after the introduction of importation cases leading to small cluster infections on household and institution level, the transmission route and the mitigating strategy are complicated and would be extended from household, institution, until population level. Monitoring and evaluating the spread of covid-19 epidemic may require the population-based model such as the susceptible-exposed-infected-removal (SEIR) model that evaluate the kinetics of susceptible, infectives, recovery, and death with and without NPIs. Such a transmission route before and after large-scale CAOs would affect the surveillance model for containing the outbreak caused by SARS-CoV-2.

There are two epidemic surveillance modes for monitoring the control of an epidemic in the absence and the presence of CAO. The former can be seen in a few countries such as Taiwan and New Zealand where there has been a lack of large-scale covid-19 CAOs for a long period before 2021 because of the strict operation of NPIs including early boarder quarantine, symptom-based screening, contact tracing, isolation, wearing masks in crowded settings, and social distancing. ^19,20^

As the epidemic surveillance model for monitoring imported cases to reduce the chance of cluster infections resulting in domestic cases would be different from that for assessing the spread of a CAO. Very few studies were conducted to provide various epidemic surveillance models in the presence and the absence of CAO during covid-19 pandemic in parallel with the evolution of SARS-CoV-2 variants.

The Taiwan covid-19 surveillance model is proposed here to demonstrate how to avert and contain covid-19 epidemics evolving with SARS-CoV-2 variants from January 2020 to August 2021, which is further divided into two phases, the non-VOC (consisting of wild type and D614G, supplementary figure 1) phase and VOC (mainly including Alpha, Beta, Gamma, and Delta VOCs, supplementary figure 1) phase. Two kinds of the epidemic surveillance models are proposed. When there was lacking of a CAO during the first non-VOC phase without high alert level of NPIs, the epidemic surveillance model, named as the transmission model from imported to domestic cases, is to monitor imported cases for detecting the resultant cluster infections of domestic cases caused by all possible transmission routes through the close contact with importation cases. In the presence of CAO during the VOC phase with high alert level of NPIs, the epidemic surveillance model with the SEIR underpinning is to evaluate whether, how, and when NPIs and vaccine, if available, can end each episode of community-acquired infection. Stemming from the coping strategies of NPIs, testing, and vaccination, the aim of this study is to present two epidemic surveillance models for monitoring and containing the spread of covid-19 caused by SARS-CoV-2 variants during the periods without and with CAOs.

**Fig 1.**
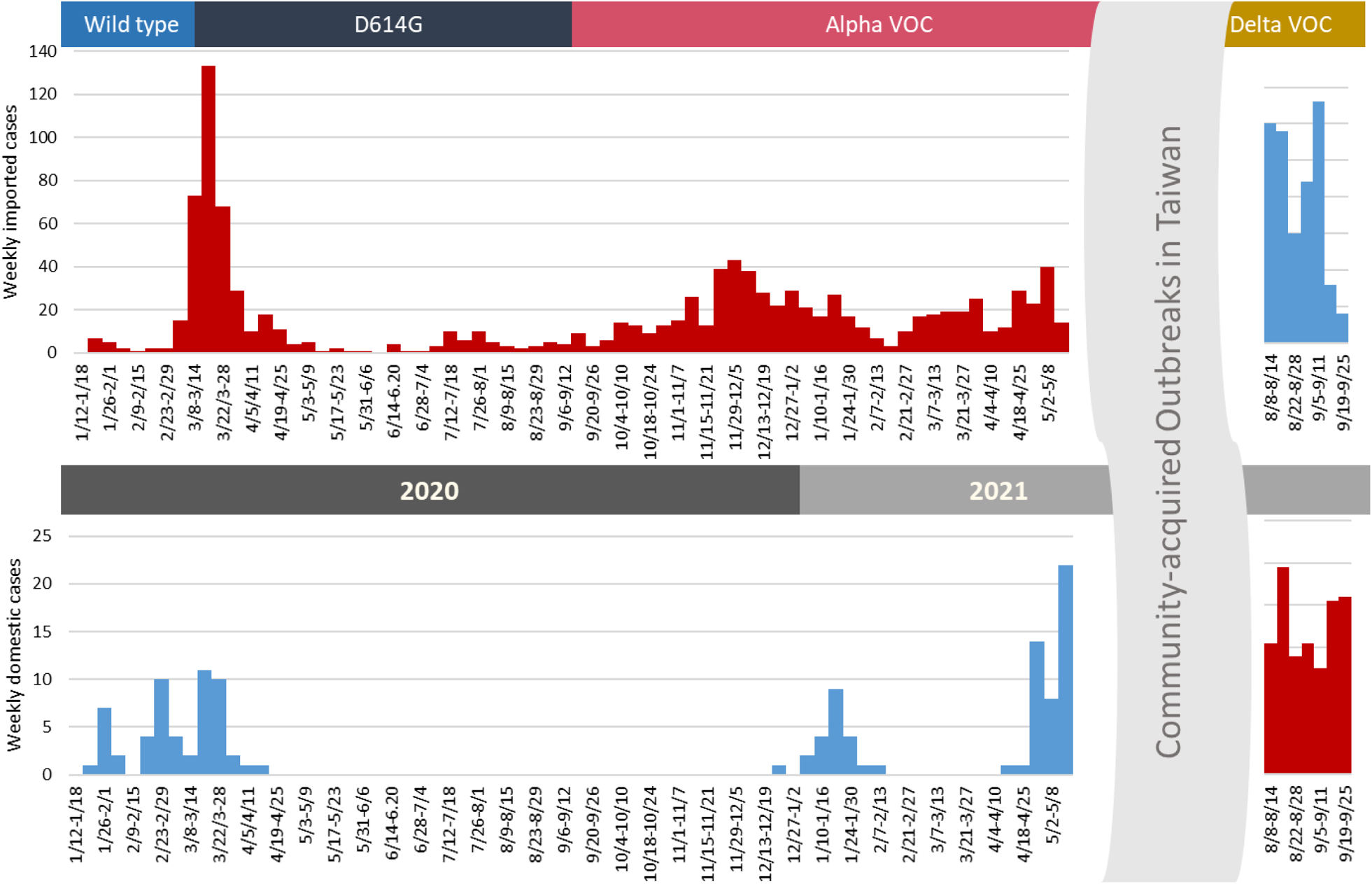
Covid-19 epidemics in Taiwan for periods without CAO by origin of cases (imported vs domestic)

## Materials and Methods

### Data source

The publicly available information on covid-19 including the daily number of cases, recovered, and deaths from 1 January 2020 to 31 July 2021 in Taiwan was extracted from the report of Central Epidemic Command Center (CECC) and the Taiwan National Infectious Disease Statistics System maintained (TNIDSS) by Taiwan Centers for Disease Control (TCDC).^21^ Regarding surveillance information system, TNIDSS has routinely collected the statistics on the number of confirmed cases of major infectious diseases, including severe complicated influenza, invasive pneumococcal disease, tuberculosis, human immunodeficiency virus (HIV), acquired immunodeficiency syndrome (AIDS), mumps, dengue fever, enterovirus infection with severe complications, Japanese encephalitis, complicated varicella, and severe pneumonia with novel pathogens.^21^ Covid-19 was categorized into severe pneumonia with novel pathogens following the first imported case confirmed on 21 January 2020.

The tabular data with epidemiologic information on covid-19 mentioned above by county and origin of cases (domestic versus imported) during the study period were obtained. The population size of 23 counties and cities in Taiwan were extracted from the official website of Department of Household Registration.^22^

### Containment measures for the non-VOC phase in Taiwan

The containment measures of the non-VOC phase in Taiwan centered on two strategies, namely border control and quarantine and isolation. They are detailed as follows. The first confirmed covid-19 case in Taiwan developed symptoms on 11 January 2020 in Wuhan and traveled to Taiwan on 21 January 2020. This imported case was quarantined at airport and later confirmed by using reverse transcription polymerase chain reaction (RT-PCR) test. Four imported cases were confirmed afterwards before the first domestic case was confirmed on 28 January 2020, who is the family contact of a previously imported case.

In response to the risk of CAO caused by the imported-domestic transmission, Taiwanese government issued a provisional regulation for border control to ban the flights from mainland China and Hong-Kong on 7 February 2020. Following the implementation of this border control regulation, there were sparse imported and domestic cases up to 5 March 2020. supplementary figure 1 shows the timelines regarding the evolution of implemented border control measures for this non-VOC phase.

There was a surge of imported cases resulting mainly from the Taiwanese returning from high risk areas abroad in mid-March, followed by an increase in domestic cases in the subsequent weeks. Fig 1 shows the number of imported and domestic cases of covid-19 by the date of onset on a weekly basis.

Following the guideline announced by Taiwan CECC, subjects with classical covid-19 symptoms including fever, cough, short of breath, fatigue, myalgia, diarrhea, and newly onset agnosia would be tested for SARS-CoV-2 infection with RT-PCR. For subjects with the history of the contact with suspected cases, it is mandatory for these subjects to be under quarantine for 14 days. The quarantined subjects were tested for SARS-CoV-2 upon the occurrence of suspected symptoms during their 14-day quarantine period.

As a supplementary measure for border control, a 14-day quarantine period is mandatory for all passengers travel to Taiwan since 19 March 2020 (14-day quarantine policy, supplementary figure 1). Subjects with classical symptoms will be tested with RT-PCR for identifying covid-19 cases during the quarantine period. The policy of testing was augmented by the requirement of a negative RT-PCT before boarding and at the end of the 14-day quarantine period since 1 December 2020.

The covid-19 cases in Taiwan in year 2020 were mainly comprised of imported cases (fig 1). The risk of CAO following the transmission of covid-19 to community caused by these imported cases were largely diminished by the border control strategies and quarantine and isolation in conjunction with NPIs such as wearing mask and social distancing.^20,23^

Supplementary table 1 lists the details on the criteria and the guidelines for the implementation of four covid-19 alert levels to stamp out CAOs in Taiwan. In brief, there is a low risk of community transmission and NPIs are least strict at the alert level one, including the requirement for wearing mask on public transportation and in crowded public venues, cancellation or postponement of no-essential gathering, and the use of social distancing (keep 1 meter apart from people in outdoor or 1.5 meter in indoor public facilities), temperature checks, and identification-based registration system for places of business and public venues. The alert level two involved with the mandatory requirement for wearing mask and social distancing at public transportation and venues with the restriction on the number of attendants under 500 and 100 for outdoor and indoor gatherings, respectively. The alert level three strengthened NPI regulation of level two to the requirement of wearing mask at all times outdoors with the restriction on the number of attendants under 10 and 5 for outdoor and indoor gatherings, respectively. Under alert level three, all places of business and public venues are closed except essential services such as medical treatment, law enforcement, and government. The alert level four involved with the lockdown, the suspension of all in-person works and schools, and the cancellation of all public events.^24^

### Containment measures for the VOC phase in Taiwan

The Alpha VOC turned in to the predominant strain of global pandemic by the end of 2020. Several outbreaks occurred in hospitals and households in Taiwan since January 2021, which evolved into a large-scale CAO in May 2021. On the top of the two strategies of border control and quarantine and isolation implemented since the non-VOC phase in Taiwan, the focus of containment measures has turned into containing CAOs by using community-based active surveillance with rapid test stations for the hotspots of outbreaks with enhanced NPIs including a strict regulation for wearing mask, stopping public gathering, setting up check points for at-risk areas such as public transportation sites and markets, and the restriction on non-essential services such as restaurants and pubs (supplementary figure 1 and supplementary table 1).^24,25^

Extended from the containment measures for the non-VOC phase in 2020 and early 2021 in Taiwan, supplementary figure 1 summarizes the timeline of the implementation of a series of containment measures for the VOC phase starting from the enhancement of NPIs from level one to level two alert following the occurrence of cases with unknown source of infection on 11 May 2021. These index cases were associated with the transmission of covid-19 occurring firstly in the hotspots in Wanhua in Taipei and the nearby townships of New Taipei City with a soaring number of covid-19 cases reported from these areas. The level three alert was thus implemented for Taipei and New Taipei City on 15 May 2021, which was rapidly extended to the nationwide level three alert on 19 May 2021 (supplementary figure 1).^24,25^

In addition to the implementation of strict NPIs by scaling up to level three alert, community-based active surveillance with rapid test stations were utilized for the identification of covid-19 cases at earliest to block the transmission in community. Since the occurrence of CAOs in early May, a series of rapid test stations were set at the hotspots of the counties and cities with outbreaks. supplementary figure 2 shows the hotspots of community-acquired covid-19 outbreak in Taiwan in the VOC phase in 2021. Among the six hotspots of CAOs in 2021, community-based active surveillance with rapid test stations were implemented in Taipei city, New Taipei City, and Miaoli county.

**Fig 2.**
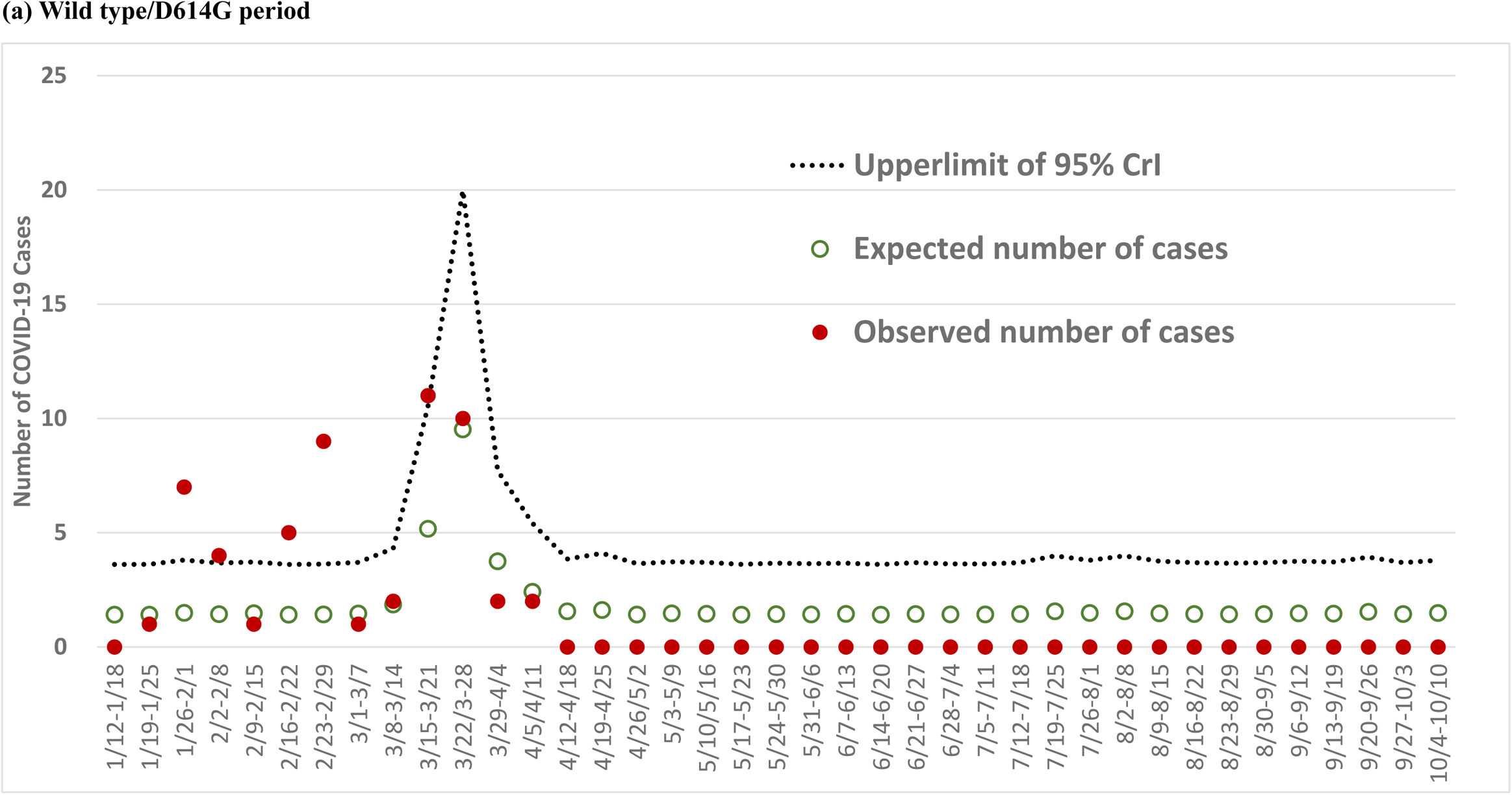
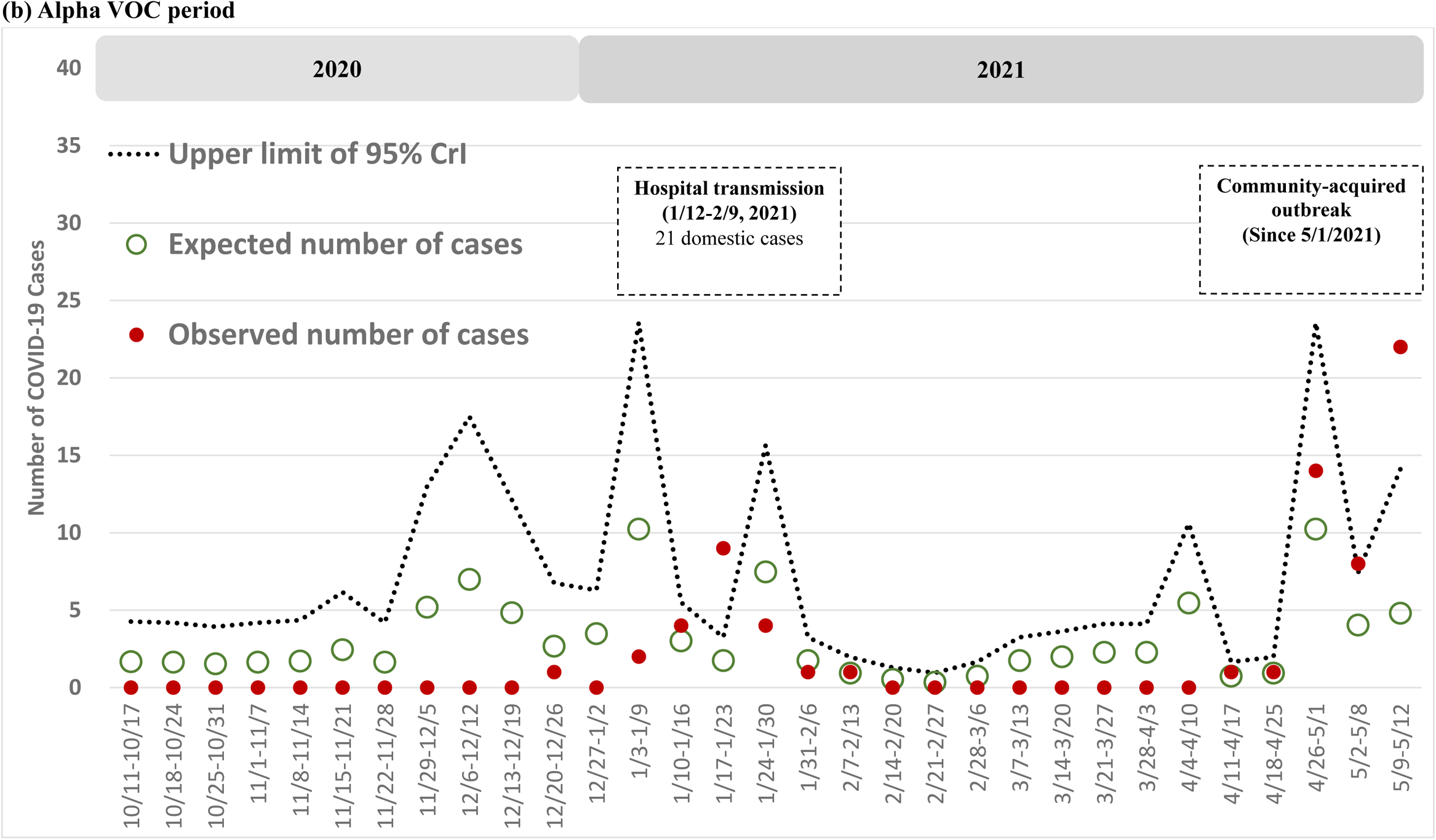
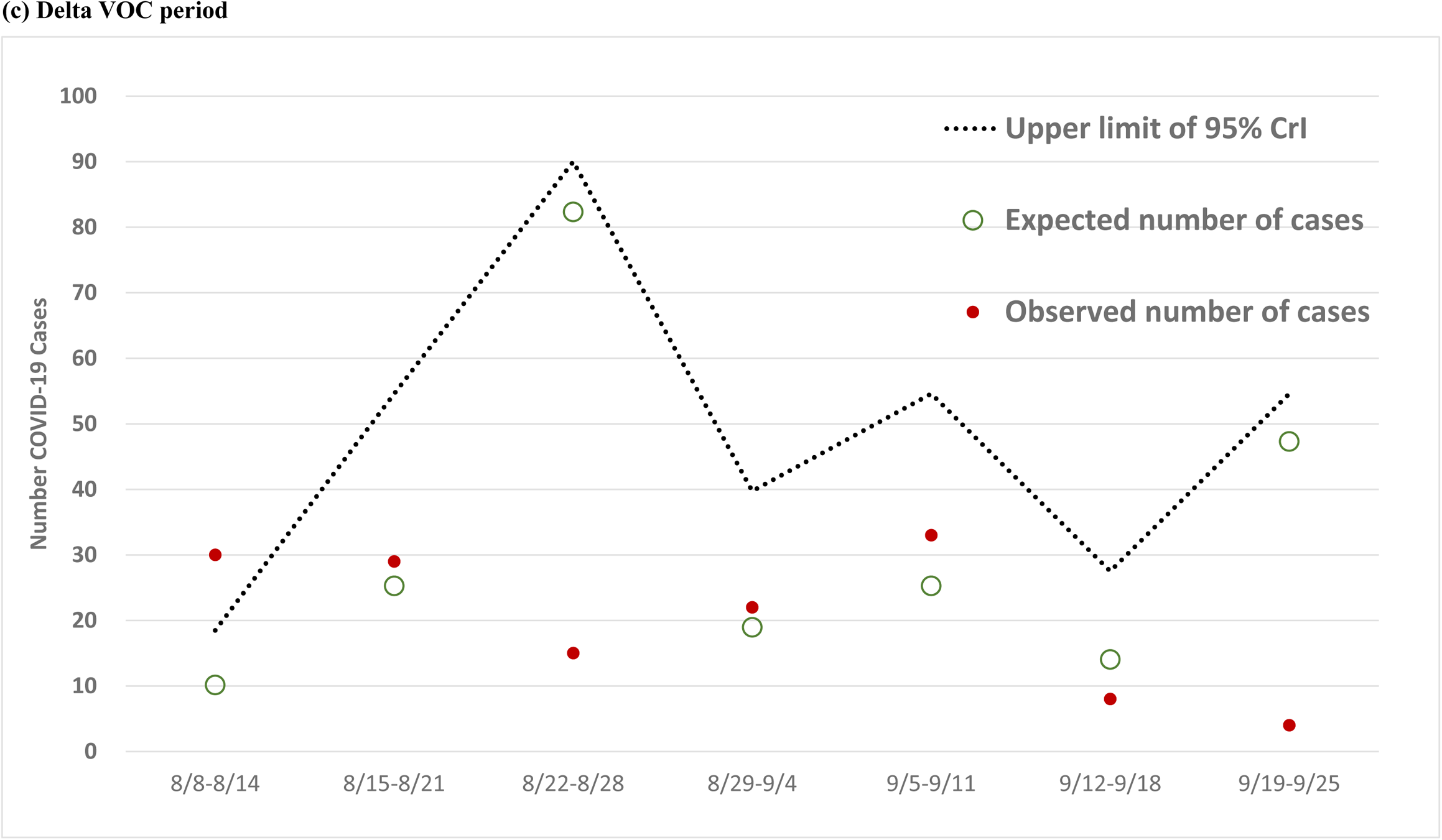
Number of observed (dotted point) and expected (green circle) domestic cases with the upper limit of 95% CrI (dotted line) by week

## Statistical analysis

### The Poisson model for assessing the risk of outbreak resulting from imported cases

We used a Bayesian random-effects Poisson regression model^26^ to calculate the expected weekly domestic infected cases following the imported cases of covid-19. The Poisson model has been widely applied to the count data on sparse cases, which occurred independently without the possibility of cluster infection. If the predicted value of our model is beyond the upper limit of 95% CrI, it means that sparse and independent assumptions based on the Poisson distribution are violated and implies the occurrence of cluster infection. The heterogeneity across cities and counties regarding the transmission of covid-19 associated with imported cases was captured by a random effect incorporated in the Poisson regression model.

Data used for estimating the parameters of the following Poisson regression model were based on imported and domestic cases between 11 January and 20 June 2020, covering the wild type and D614G period in Taiwan. Because of the imported cases would take incubation time to generate secondary cases in communities, we tested the lag time of imported cases by zero (concurrent) week, one week, and two weeks, and further selected the optimal lag time interval with the smallest deviance information criterion (DIC). The Bayesian Markov chain Monte Carlo (MCMC) method with non-informative priors was used to estimate the posterior distribution of parameters including the risk of imported-domestic transmission and the county-specific random effect. The posterior samples were generated by using 250 000 simulations following the burn-in period of 100 000 iterations with the thinning interval of 10. On the basis of the 25 000 MCMC posterior samples, the mean values and the 95% CrI can be derived accordingly. The one-week lag model was used for evaluation on the basis of the lowest DIC (255.8) compared with that of the concurrent-week model (260.3) and two-week lag model (279.5) for imported cases.

We used the upper limit of 95% CrI generated by the parameters updated by the data on the non-VOC phase in Taiwan as the threshold for assessing whether cluster infections following imported cases resulted in an emerging CAO. We also used this threshold for alerting the possibility of yielding a large-scale CAO through the imported-domestic transmission in the subsequent week. The possibility of CAO was deemed low if observed domestic cases were not more than the upper limit of the 95% CrI. Otherwise, a CAO was likely to occur and therefore would warrant the implementation of relevant containment measures to prevent a community outbreak.

Regarding the impact of imported cases on the occurrence of domestic cases for the Alpha (11 October 2020 to 12 May 2021) and Delta (8 August to 18 September 2021) VOC phase without CAOs in Taiwan, a Bayesian Negative Binomial regression model was applied to taking into account the cluster effect associated with the transmission of covid-19. Following the approach applied for the wild type and D614G period, the one-week lag model was adopted. The estimated results on the risk of imported-domestic transmission and the upper limit of the 95% CrI of expected domestic cases were used for the surveillance of covid-19 in the wake of the second wave of covid-19 pandemic in October 2020 and the Delta VOC period following the stamping out of CAOs during May to July 2021 in Taiwan.

### Assessing the efficacy of containment measures for the VOC phase in Taiwan

The CAOs in the VOC phase in Taiwan were characterized by using the overall and area-specific incidence between 1 May and 31 July 2021. The data used for analysis consisted of covid-19 cases reported by TCDC during this period.^21^ The area-specific incidence of covid-19 was calculated by dividing the total number of daily cases of each county and city by the population at risk in each area. For a better presentation of the trend of covid-19 incidence, a moving average with a three-day window was applied to avoid the influence of daily fluctuations. By comparing the trend of covid-19 incidences between areas, the effect of containment measures with and without a mass screening campaign were assessed.

The extents of NPIs and testing for containing the community-acquired covid-19 outbreaks were assessed by using a time-varying transmission coefficient estimated from the standard compartment model for an infectious disease. Stemming from the method elaborated by Daley and Gani,^27^ the transmission coefficient was estimated by using the empirical information on the frequencies of susceptible (S), exposed (E), infected (I), and removal (R, including recovery and death) subjects in conjunction with a compartment model depicting the dynamic of disease status of a population. While the two rates determine the evolution of covid-19 to the state of infective and recovered were assumed time-invariant, a time-varying transmission coefficient during the period of outbreak captures the change of covid-19 transmission in community affected by the implementation of NPIs and testing. The value of 0.2 and 0.14 were adopted for the two time-invariant rates, σ and α, respectively, by using the information reported in previous studies.^28,29^ The effectiveness of the containment measures for the VOC phase can thus be estimated by

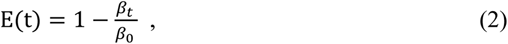

where β_0_ and β_t_ represents the estimated results on the transmission coefficient at initial period and the *t*^th^ epoch since the occurrence of CAOs in May 2021 in Taiwan. The effective reproductive number, R_t_, can then be derived from the estimated results on the time-varying transmission coefficient, β_t_.

### External validation

To validate the proposed surveillance model for the transmission model from imported to domestic cases during the non-VOC period in Taiwan, the publicly available covid-19 data provided by the Ministry of Health in New Zealand were used. The first covid-19 was reported in New Zealand on 26 February 2020.^30^ Data on age, sex, District Health Bureau, travel overseas, flight details, and arrival date, when applicable, were extracted from 23 February 2020 to 28 February 2021.

Regarding the validation for the surveillance model for the period with CAOs, the data on three hotspots of the Alpha VOC outbreak occurred in Taiwan between May and July 2021 were used. The two hotspots in Northern Taiwan, namely Taipei and New Taipei City represent the first wave of the CAO with the initial phase of scaling up NPIs and the rolling out of testing. The hotspot in Central Taiwan, Changhua county is the second wave of this CAO with cluster infections introduced by the communication with the areas of first wave CAO. The chronological order of the incidence of covid-19 for these hotspots were compared to validate the epidemic surveillance model for CAO.

### Patient and public involvement

Patients were not included in the design or conduct of this study.

## Result

### The Surveillance Model for Cluster Infections Transmitted from Imported Cases

Fig 2 shows the weekly domestic cases of the observed number (red dot) and the expected number (green circle) for the wild type and D614G period (January to September 2020; fig 2 (a)), the Alpha VOC period (October 2020 to May 2021; fig 2 (b)) before the outbreak of CAOs in Taiwan in mid-May 2021, and the Delta VOC period (mid-August to September 2021; fig 2 (c)) after containing CAOs. Table 1 shows the details of the estimated results of the parameters encoded in the Poisson regression model with a one-week lag of imported cases regarding the three periods without CAO in Taiwan, namely the wild type and D614G period, Alpha VOC period, and Delta VOC period.

**Table 1.** Estimated results for the risk of imported-domestic transmission of covid-19 for the three periods in Taiwan.

| **Parameter** | **Estimate** | **95% CI** |
| --- | --- | --- |
| ***Wild type/D614G period***<br>**(January to September 2020)** |  |  |
| Common intercept | -3.5457 | -5.1978 to -2.4413 |
| Risk of imported-domestic transmission | 0.0954 | 0.0644 to 0.1259 |
| Standard error of random intercept, $\sigma_v$ | 1.8116 | 0.9861 to 3.7359 |
| ***Alpha VOC period***<br>**(October 2020 to May 2021)** |  |  |
| Intercept | -1.9448 | -4.1238 to 0.0712 |
| Risk of imported-domestic transmission | 0.1414 | 0.0541 to 0.2510 |
| Dispersion parameter | 1.7438 | 0.2734 to 3.8218 |
| ***Delta VOC period***<br>**(August to September 2021)** |  |  |
| Risk of imported-domestic transmission | 0.1016 | 0.0701 to 0.1359 |
| Dispersion parameter | 1.4851 | 0.3664 to 3.2449 |

The upper bound of 95% CrI of those expected cases (dotted line, fig 2) is plotted on the basis of the Poisson regression model in order to provide the threshold of a cluster infection in community caused by the transmission from imported cases one week before. This one-week prior alert on the risk of elevated imported-domestic transmission raises the vigilance on NPIs for averting further CAOs.

#### Wild Type and D614G period

During the wild type/D614G period, the estimated results based on the Poisson random-effect regression model revealed that an imported case was at greater risk for the ensuing cluster infections by 9.54% (95% CrI: 6.44 to 12.59%, table 1). Fig 2 (a) shows that there were five weeks (26 January to 1 February, 2 February to 8 February, 16 February to 22 February, 23 February to 29 February, and 15 March to 21 March) in which the observed numbers of domestic cases exceeded the upper bound of the 95% CrIs of expected cases. This period yielded 81% of clustered cases (N=22 of 27 community-acquired cases) in five clusters in Taiwan, including three household clusters (with five, three, and six covid-19 cases, respectively), a medical institute cluster (with nine covid-19 cases), and one academic institute cluster (with four covid-19 cases). Learning from this early period of the wild type of covid-19 provides a strong rationale for being on alert for the ensuing cluster infections preceding one week when imported cases were introduced. This accounted for why none of these five cluster events led to any large-scale CAO in Taiwan because of the advocacy of strict NPIs through CECC.

Fig 2(a) shows such a surveillance model was very useful particularly when there was a substantial surge in imported cases resulting from large-scale covid-19 pandemic worldwide. This can be seen in our cases as shown in fig 1 between March and April 2020. Again, the surveillance model was used for alerting the possible cluster infections to stop further community-acquired outbreak. Alerted by the upper bound of 95% CrI for the expected domestic cases (20 cases) and the expected domestic cases at weekly basis, the observed domestic cases were kept lower than the upper limit of 95% CrI to avoid large-scale CAOs in April. Since then, there has not been any domestic case until December 2020.

#### Alpha VOC period

There has been no CAO during early Alpha VOC phase of covid-19 pandemic from Oct. to Dec., 2020. October to December 2020. The second surge of imported cases occurred from January 2021 onwards (fig 1). Fig 2 (b) shows the observed and predicted domestic cases as a results of imported-domestic transmission with the upper limit of 95% CrI overlaid as the upper bound. Again, there was one week, 17-23 January 2021 in which the observed numbers of domestic cases were beyond the upper bound of surveillance level resulting from hospital-based cluster infections and three subsequent household clusters (11 family members). The source of this cluster infection was later identified as an imported case infected by Alpha VOC. After being alerted by the proposed surveillance model and the timely containment measures, including quarantine to all staffs in the hospital and their family close contact for 14 days, there was no further CAO until early May 2021 when the observed domestics case reached beyond 20 that had far beyond the expected surveillance curve (5 cases) and the corresponding upper bound of 95% CrI (14 cases). The estimated results on the basis of the empirical data during this Alpha VOC period shows an increase in an imported case was at greater risk for the ensuing cluster infections by 14.14% (95% CrI: 5.41 to 25.10%, table 1). The ensuing community-acquired outbreak on Alpha VOC was noted and described in a separate section below. This outbreak has lasted for two and half months and subsided until July 2021.

Generally speaking, the epidemic surveillance model for monitoring the ensuing cluster infections in the wake of imported cases was demonstrated through three periods from the wild type, D614G, until Alpha VOC. The upper bound of 95% CrI for expected domestic cases derived from the imported cases one week prior provide a good guidance for the threshold of CAO in each country or region when the surge epidemic of covid-19 has restarted from the introduction of imported cases. Such a threshold value would be affected by the underlying coverage rate of vaccination and the extent of NPIs.

In Taiwan scenario shown here, the threshold value for weekly domestic cases during three SARS-CoV-2 variants would not beyond 20 cases under the low coverage rate of vaccination and good performance of NPIs. The peak of domestic cases far beyond the threshold vale in early May 2021 not only presaged the ensuing CAOs as below but also revealed the loose of NPIs at that time in Taiwan.

### The Epidemic Surveillance Model for Monitoring Community-acquired Outbreak

#### Epidemiologic characteristics of the second phase with community-acquired outbreak in Taiwan

In Taiwan, it did not have a large CAO until mid-May 2021 although there had been a series of episodes on cluster infections far beyond the threshold of the expected epidemic curve as depicted in early May of fig 2 (b). The VOC phase of covid-19 outbreak occurred in Taiwan after mid-May starting from a series of clustered events. From 11 May 2021, the community outbreak emerged in Taiwan heralded by six domestic cases with unknown infection source. Following this clustered event, a nationwide level three alert was announced on 11 May (supplementary figure 1). Characteristics of epidemic curves after this CAO are described as follows.

#### Incidence trend by three categories, hotspots and cluster infections

There was a CAO emerging from Wanhua, one of the districts with a lower socioeconomic status in Taipei, and spreading across other surrounding districts of Taipei, and also other counties and cities of Taiwan including New Taipei City, Taoyuan, Miaoli, Changhua, and Keelung. Fig 3 shows the daily number of covid-19 cases for this CAO and the subsequent episodes. Fig 3 (a) and (b) show the epidemic curve and the corresponding incidence trend of covid-19 in Taiwan from outbreak until subsidization. It took around two and half months after scaling up level two to nationwide level three of NPIs together with community-based active surveillance of rapid test stations beginning from mid-May. Notably, Fig 3 (b) shows the time trend of covid-19 incidence in Taiwan associated with the implementation of community-based active surveillance of rapid test stations. By using the active surveillance of the hotspots and its neighborhood, the covid-19 cases, with or without clinical symptoms, can be identified at earliest. The peaks of incidence trend corresponding to the setting of rapid test stations revealed this effect.

**Fig 3.**
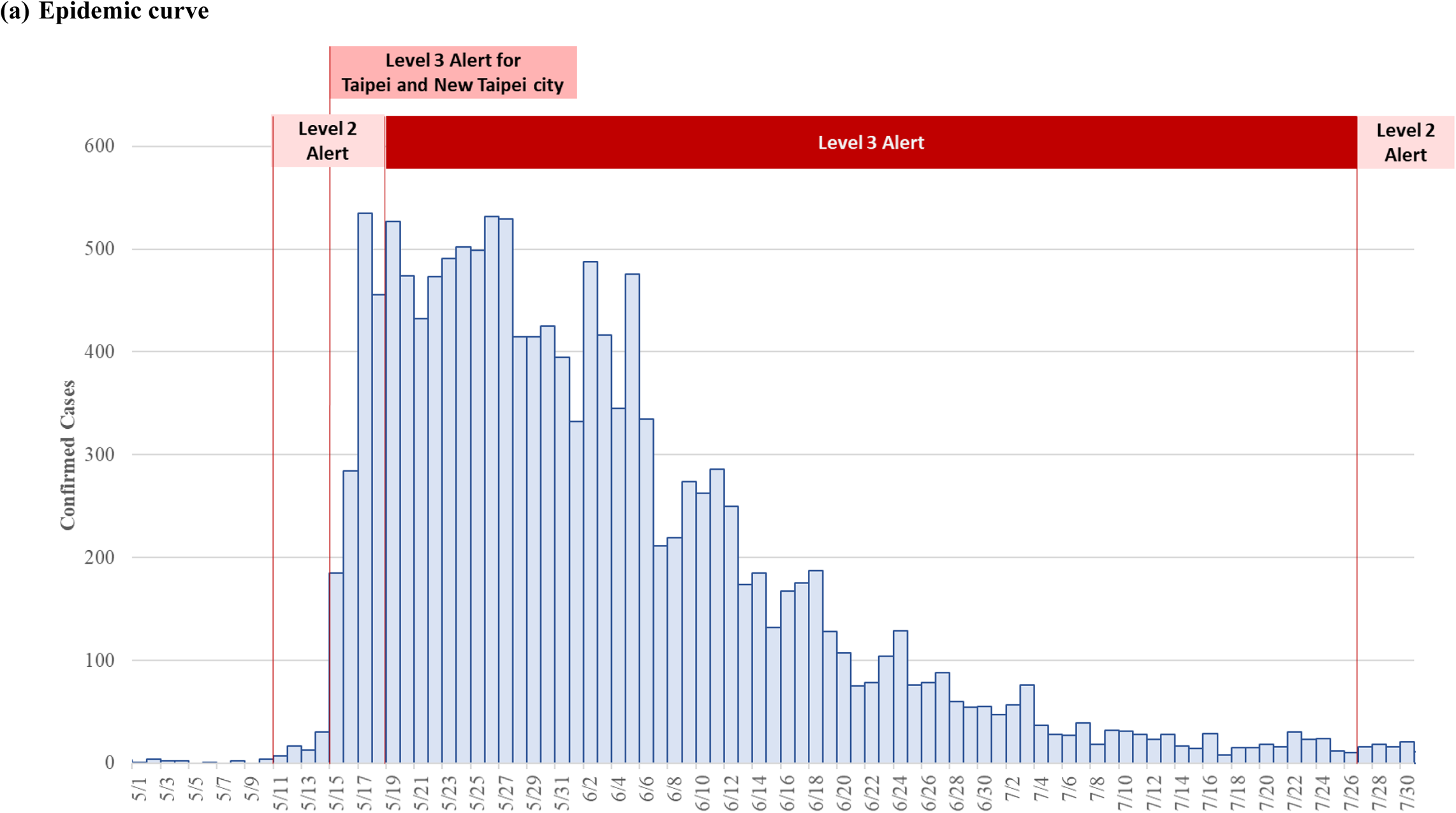
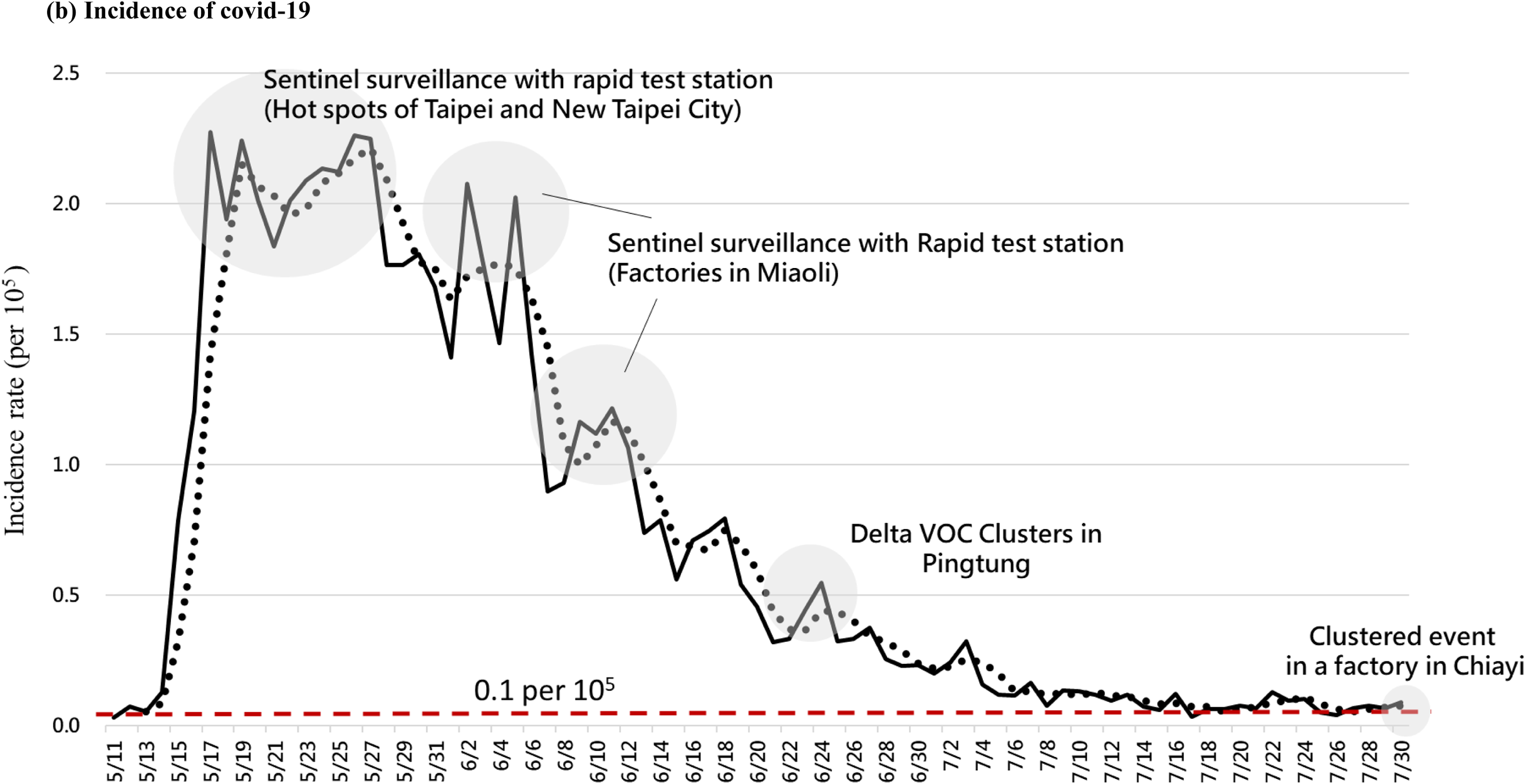
The epidemic curve of community-acquired covid-19 outbreak in May 2021, Taiwan

supplementary figure 3 shows the incidence trends of six hotspots from the orifice of the emerging outbreak at Taipei with highest incidence to the second wave of outbreaks with lower incidence. The latter may be reduced by the introduction of level three alert of NPI after mid-May. Based on these findings, county-specific incidence rates in Taiwan can be categorized into high, medium, and low areas during CAOs. In addition to the transmission of covid-19 in the community, the high incidence areas were mainly because of early detection of covid-19 cases after transmission via several community-based active surveillance of rapid test stations including three earlier hotspots such as Taipei, New Taipei City, and Miaoli county.

The incidence rates reached the peak of around 10 per 100 000 as shown in supplementary figure 3(a). Counties in the medium incidence areas represented the second wave of outbreaks through the transmission route of two earlier hotspots from Taipei and New Taipei City. The peaks for the incidence rate resulting from these cluster infections was 2.2 (per 100 000) for the fruit merchant cluster in Changhua, 1.3 and 4.9 (per 100 000) for the household clusters in Taoyuan and Keelung, and 4.6, 4.3 and 2.5 (per 100 000) for restaurant, long-term care facilities, and nursing home clusters in Keelung (supplementary figure 3 (b)). The incidence rates for other counties were extremely low due to the implementation of level three alert of NPI as shown in supplementary figure 3 (c).

#### R_t_ and effectiveness of NPIs and testing in Taiwan

Fig 4 shows the estimated results on the effectiveness of NPIs and testing implemented in Taiwan during the VOC phase. After the implementation of the level three alert, the containment measures including NPIs and testing effectively curbed the outbreak. The R_t_ decreased from 4.0 to 0.7 from 18 May 2021 to 31 July 2021. The effectiveness of NPIs and testing which reflects the strategies implemented two weeks ago was over 60% after 26 May 2021 and enhanced to over 90% after 14 June 2021. Supplementary table 2 lists the estimated results on the time-varying transmission coefficient (β_t_), effective reproductive number (R_t_), and the effectiveness of NPIs for each epoch derived from the empirical data on the covid-19 outbreak in Taiwan during the Alpha VOC period in 2021.

**Fig 4.**
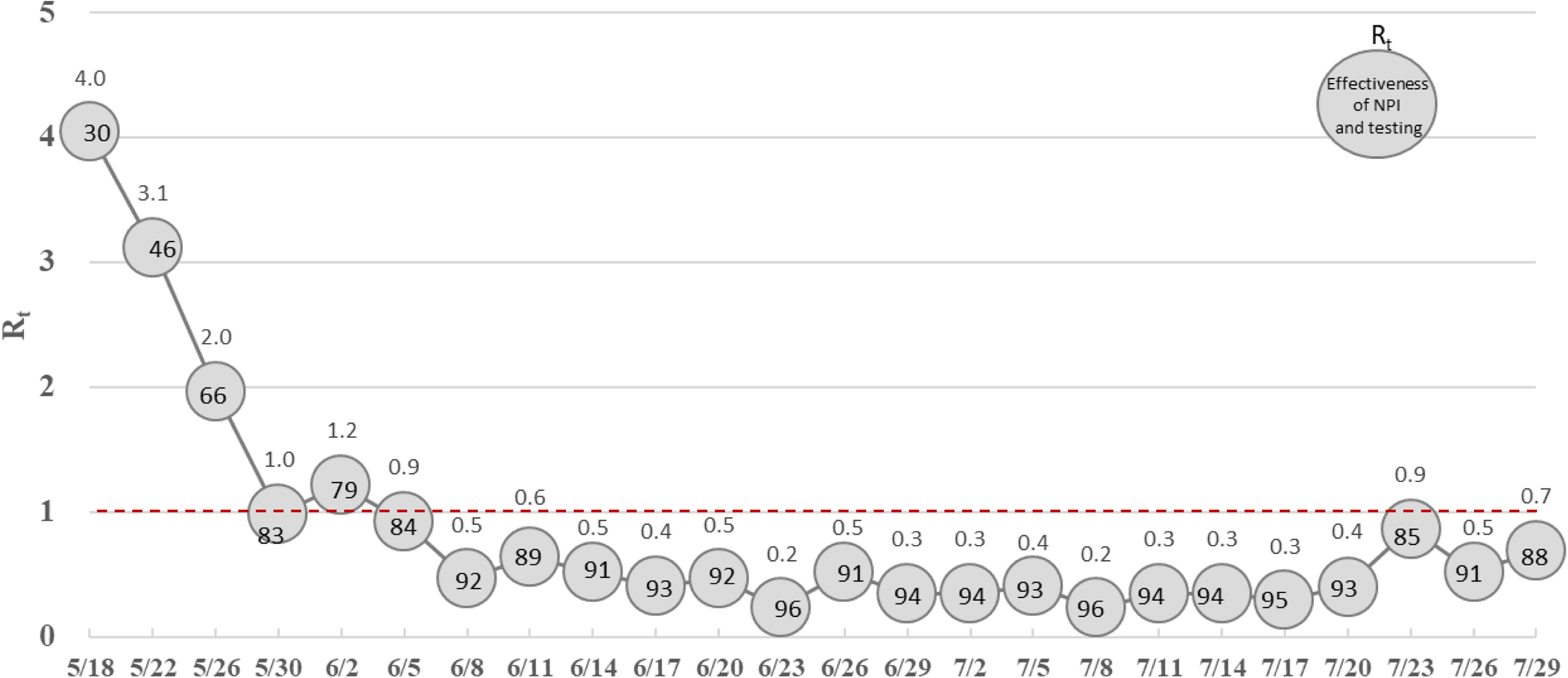
R_t_ and effectiveness of NPIs and testing in Taiwan

#### Containing Delta VOC Cluster Infection in Pingtung, Southern Taiwan

During the VOC phase of covid-19 outbreak in Taiwan, Delta variant also caused a cluster infection in Pingtung, a county in Southern Taiwan. Starting with an index case confirmed on 11 June, this clustered event spanning 25 days evolved rapidly to the contacts of the index case including neighbors, driver, household, and friends (supplementary figure 4), resulting in a total of 17 cases in this outbreak. supplementary figure 5 (a) shows the epidemic curve of the outbreak from 11 June to 5 July. After the Delta VOC was identified as the viral strain responsible for this clustered event, the enhanced containment measures including community-based active surveillance with rapid test stations and quarantine and isolation were implemented in Pingtung since 23 June. The R_t_ for the initial period (11-22 June 2021) and later period (23 June to 5 July 2021) were estimated as 3.2 and 0.3, respectively, suggesting an effective containment of this outbreak resulting from Delta VOC in Pingtung.

**Fig 5.**
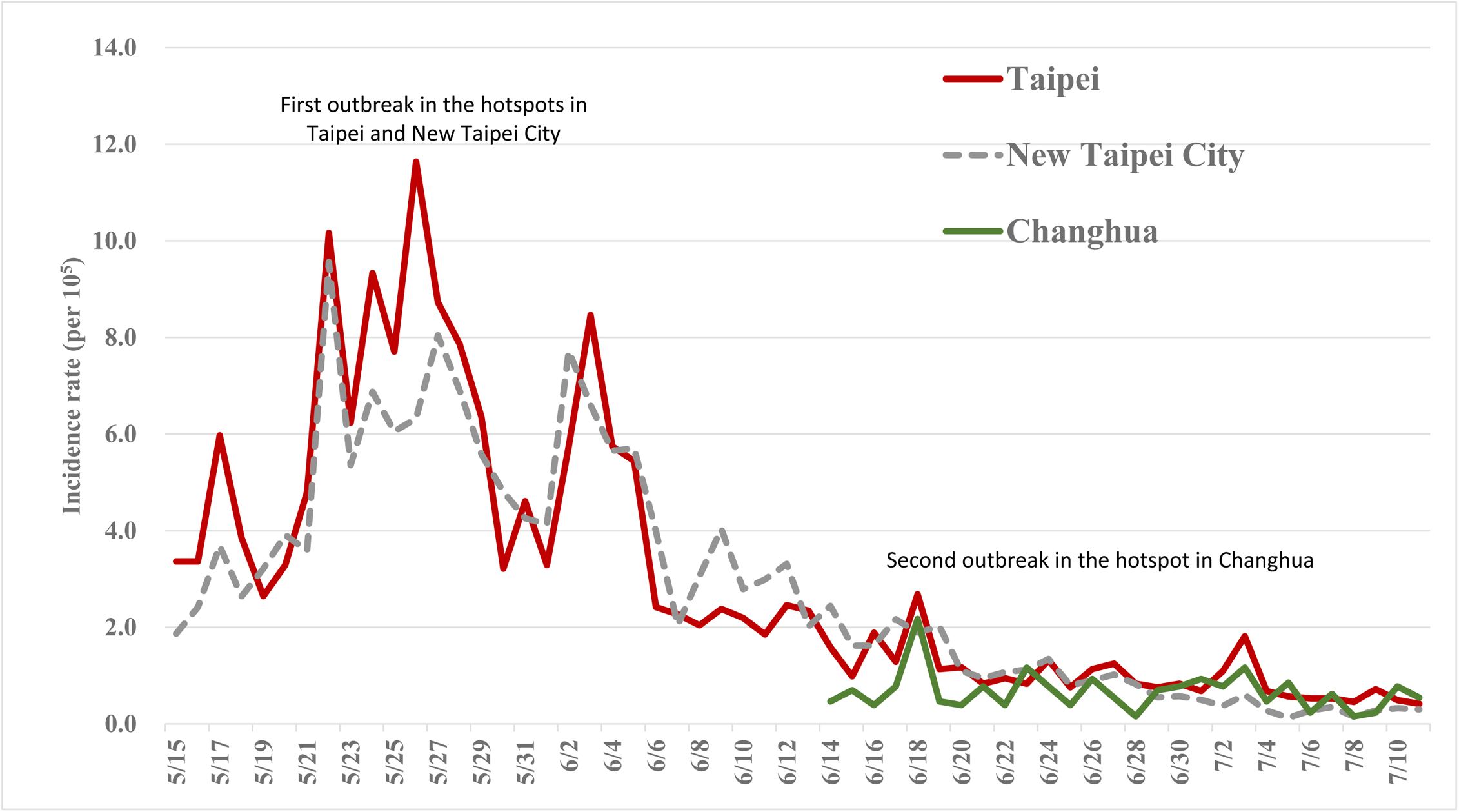
Covid-19 incidences in the two hotspots of the first outbreaks in Taipei and New Taipei City and that of the second outbreak in Changhua.

To assess the effectiveness of NPIs and testing exerted in Pingtung for containing the CAO associated with Delta VOC, the transmission coefficient and R_0_ of 1.09 and 7.6, respectively, were applied (Campbell et al., 2021). Driven by this force of transmission and assuming a low level of NPIs in Pingtung (30%, as of the estimated results for 16-18 May in Taiwan, fig 4), the expected number of cases and epidemic curve caused by Delta VOC without the implementation of enhanced containment measures were projected (supplementary figure 5 (b)). Had there been no effective containment measures, a total of 75 cases would be expected, which gives the estimated results on effectiveness of NPIs and testing in reducing covid-19 cases by 77%.

#### Return of Imported-domestic transmission for Delta VOC Period

After controlling the CAO, it returned to monitor the imported-domestic transmission mode for Delta VOC. The Poisson-based epidemic surveillance model was applied to monitoring imported cases dominated by Delta VOC. The estimated results (table 1, Delta VOC period) show an increase in an imported case was at greater risk for the ensuing cluster infections by 10.16% (95% CrI 7.01% to 13.59%). Fig 2 (c) shows the observed and expected number of domestic cases along with the upper limit of 95% CrI projected from the estimated results of table 1. Although there were two weeks (22-28 August 2021 and 19-25 September 2021) by when the surge in the number of domestic cases as a result of imported-domestic transmission were expected (green circle), the observed number of domestic cases is far below the threshold of outbreak. This can be attributed to the implementation of enhanced containment measures including the border control strategies strengthened by multiple tests on arrival and during quarantine, the collective quarantine strategy, and the elevated alerts of NPIs to level two and three since the CAO in May 2021 in Taiwan.

### External Validation of the Epidemic Surveillance Model for Importation Cases

Supplementary table 3 shows the estimated result by applying the Poisson surveillance to the data on New Zealand covid-19 outbreak in 2020. Notably, for each imported case in New Zealand, the risk of CAO was increased by 9.38% (95% CrI 8.88%-9.86%), a figure close to the estimated results for Taiwan (9.54%, 95% CrI 6.44% to 12.59%). Details regarding the spatial temporal distribution of COVID-19 outbreak by types of cases in New Zealand are provided in supplementary figure 6.

Supplementary figure 7 shows the expected number of cases projected from the Poisson model by using the parameters trained from the empirical data of New Zealand (supplementary table 3). Similar to the application in Taiwan, the risk of CAO associated with imported cases can be assessed by comparing the observed cases (red dot in supplementary figure 7) with the upper limit of the 95% CrI of expected cases (dotted line in supplementary figure 7). Supplementary material C gives the detailed interpretation on the result of this external validation.

### External validation of the Epidemic Surveillance Model for Community-acquired Outbreak

Fig 5 presents the incidences of the first CAO took place in two hotspots of Taipei and New Taipei City and that of the second wave of the hotspot in Changhua. It clearly shows the force of transmission from the first outbreak in Northern Taiwan (Taipei and New Taipei City) were contained in the outbreak occurring in the hotspot in Middle Taiwan (Changhua) resulting from the implementation of level three alert of NPIs.

## Discussion

While a series of cyclic CAOs in each country or region during covid-19 pandemic have been observed since 2020, it is imperative to avert such cyclic CAOs by developing the epidemic surveillance models for forestalling covid-19 epidemic through monitoring the possible spread of imported cases and for containing the spread of SARS-CoV-2 in the face of CAOs. By using the empirical data on imported and domestic covid-19 cases in Taiwan, the risk of weekly domestic cases resulting from one-week prior imported cases were estimated as 9.54% (95% CrI 6.44% to 12.59%), 14.14% (95% CrI 5.41% to 25.10%), and 10.16% (95% CrI 7.01% to 13.59%) for the wild type and D614G, Alpha VOC, and Delta VOC, respectively. A one-week-prior warning by using the upper bound of 95% CrI of the imported-domestic transmission were signaled to avert the resultant CAO based on these estimated results. Regarding the real time monitoring for the containment of CAOs, NPIs strengthened from 30% on 18 May, 2021 at the initial outbreak in Taiwan to more than 90% since 8 June were revealed, which contributed to the reduction of R_t_ form four to lower than one since then. The merit and the application of these two epidemic models are described as follows.

### Averting CAO caused by imported-domestic transmission model

Applying the first surveillance model for monitoring importation cases prior to one week of forming cluster infections of domestic covid-19 cases is very useful for alerting the surrounding community in proximity to that of importation cases to enhance NPIs and active rapid test with contact tracing for a high value of expected cases or the observed cases exceed the threshold of the upper bound of 95% CrI. This accounts for why there has been no CAO before mid-May in Taiwan. Such a good control over covid-19 epidemic has been reported by the previous studies by evaluating NPIs on individual level and population level^10,20,23^ and also using the duration from R_t_ larger than one to R_t_ smaller than one and case load following the machining learning model.^33^ The consistent results in terms of the risk of CAO caused by each imported case illustrated in Taiwan and the external validation in New Zealand further add the credibility to the application of this surveillance model in the scenario without CAO.

Applying the proposed imported-domestic transmission model, five clusters with a potential of CAO were identified by presaging the expected number of domestic cases beyond the threshold of CAO based on one-week prior imported cases and further providing alert message for stringent local NPIs and active rapid testing with contact tracing. The CAO was therefore averted during the wild type and D614G period. In contrast, large-scale CAO could not be averted during the Alpha VOC period while the expected number of domestic cases was far beyond the threshold of CAO between 9-12 May because of the increased transmissibility of Alpha VOC that was supported by the increased risk of imported-domestic transmission in comparison with the wild type and the D614G type (14.14% vs 9.54%, table 1). However, low level of NPIs might also make contribution to such a CAO around mid-May (40%, fig 4). Regarding the Delta VOC period after stamping out CAO in relation to Alpha VOC in Taiwan, the level of NPI alert and the strict border control strategies implemented since the CAO period reduced the risk of imported-domestic transmission to 10.16% (table 1) to avert Delta VOC CAO.

### Stamping out CAOs with real-time epidemic surveillance model

The application of the second SEIR model for monitoring the real-time operation of NPIs after scaling up alert provides a clue to whether, how, and when to contain CAOs with efficiency, particularly for VOCs. This is especially true for the illustration in Taiwan, since the mass vaccination was at the initial stage of rolling out period with an insufficient vaccine supply before and during the CAO in May 2021 in Taiwan. The CAO in relation to Alpha VOC in Taiwan was stamped out effectively through the implementation of NPIs by scaling up the nationwide alert level and the establishment of active surveillance at hotspots (supplementary figure 1 and figure 3). The real time monitoring for the implementation of NPIs can be also quantified by using the SEIR-based surveillance model (fig 4). In contrast to Taiwan, Israel demonstrated the effective containment of CAO by using mass vaccination.^31,32^ The impact of these two approaches, NPIs with testing and vaccination, on stamping out CAOs can be assessed by using the proposed SEIR-surveillance model for a real-time informed decision making to gauge the level of NPI required in the face with large scale CAO. Similar situation may be encountered in the face of the emergence of VOCs particularly with high risk of vaccine breakthrough. This also account for why it only took two and half of months in the illustration of Taiwan scenario to curb Alpha and Delta covid-19 epidemic mainly because of strict operation of NPIs on population level together with community-based active surveillance of rapid test and partially because of 40% coverage of one-dose vaccine.

### Applications of the imported-domestic transmission surveillance model for averting resultant CAOs

The first kind of surveillance model for imported-domestic transmission of covid-19 is to reduce domestic cases through the possible transmission via close contact with imported cases by means of effective quarantine and isolation or NPIs. In addition to the illustration of Taiwan experience, this model can be also applied to those countries with the controllable community-acquired outbreak such as Israel^34^ and Qatar^35^ after the mass vaccination program since early 2021 to monitor the impact of imported cases on the risk of domestic cluster infections. This is especially important for CAOs resulting from vaccine breakthrough in the country or region with high vaccine coverage or a well-controlled covid-19 attributed to strict NPIs reported worldwide. The former situation is noted in the recent Delta VOC outbreaks of Singapore^37^ and Israel^38^ after mass vaccination and the latter can refer to the classical example of Australia^39^ with a well-controlled covid-19 via high level NPI alert for a long time.

### Real-time monitoring of CAO for informed decision making on containment measures

The second kind of surveillance model for CAO is to monitor whether, how, and when NPIs and community-based active surveillance with rapid test are effective in containing CAO. This surveillance model for CAO is very useful for countries that have lasting CAO and low supply of vaccine and rely mainly on NPIs as the main containment strategy. Namely, countries at their rolling out stage to raise vaccination coverage require such a support from the real-time surveillance of CAO to guide the implementation of appropriate NPIs to mitigate CAO with the balance of socioeconomical activities.

There are two major circumstances that require the refinement of the current proposed epidemic surveillance models. These two models have not incorporated health care capacity for accommodating the threshold of tolerable covid-19 cases responsible for each episode of CAO. In the consideration of resuming pre-pandemic activity, making allowance for this factor becomes paramount important for the implementation of NPIs and testing given vaccine coverage rate. Different countries and regions may require different thresholds of CAO based on two models. With the observation on increasing cases with vaccine breakthrough, the rapid emergence of VOCs with a wide spectrum of immunogenicity, high transmissibility, and resistance to the antibody caused by natural infection or vaccination, and the waning immunity among the population, there is a high likelihood for the establishment of SARS-CoV-2 in population.^36^ Given the possibility of this long-term association of SARS-CoV-2 with human population, the goal of epidemic surveillance may shift from the elimination of this pathogen to the balance between healthcare capacity, socioeconomical activity, and population immunity. If it is such a case, the proposed surveillance models should take this factor into account and then can be used as a guide to inform the containment measures required to mitigate large scale CAO bounded by the healthcare capacity. Second, as the border control policy on quarantine and isolation of importation cases would be altered with the advent of high performance of rapid test and the gradual expansion of vaccine coverage rate worldwide, the first kind of surveillance model for monitoring imported-domestic transmission to avert CAO may vary from country to country, depending on the extent of NPIs, the administration of test, and the coverage rate of vaccine. Such a heterogeneity should be taken into account to refine the surveillance model on imported-domestic transmission when it is applied to averting a large-scale CAO.

In conclusion, two kinds of epidemic surveillance model are proposed and also applied to containing SARS-CoV-2 variants from non-VOC phase to the VOC phase based on Taiwan scenario. These two models can be accommodated to monitor the epidemic of forthcoming emerging SARS-CoV-2 VOCs with various circumstances of vaccine coverage, NPIs, and test in countries worldwide.

## Contributors

AMY, THC, and SLC conceptualized and design the study. AMY was responsible for data analysis and the drafting of the manuscript. WJC, TYL,GHJ and CYH was responsible for statistical analysis. WJC, TYL, and GHJ were in charge of the data collection and management. THC, CYH, STW, HD and SLC interpreted results and revised the manuscript. All authors agreed the findings and provided input on the revision of the manuscript.

## Ethical approval

This study uses publicly available case line list without any private and identifiable information and does not require IRB approval.

## Data sharing

All data are available on request.

## Data Availability

All data are available on request.

## Acknowledgments

Not applicable.

## Supplementary material

### A. Containment measures implemented in Taiwan

**Supplementary figure 1.**
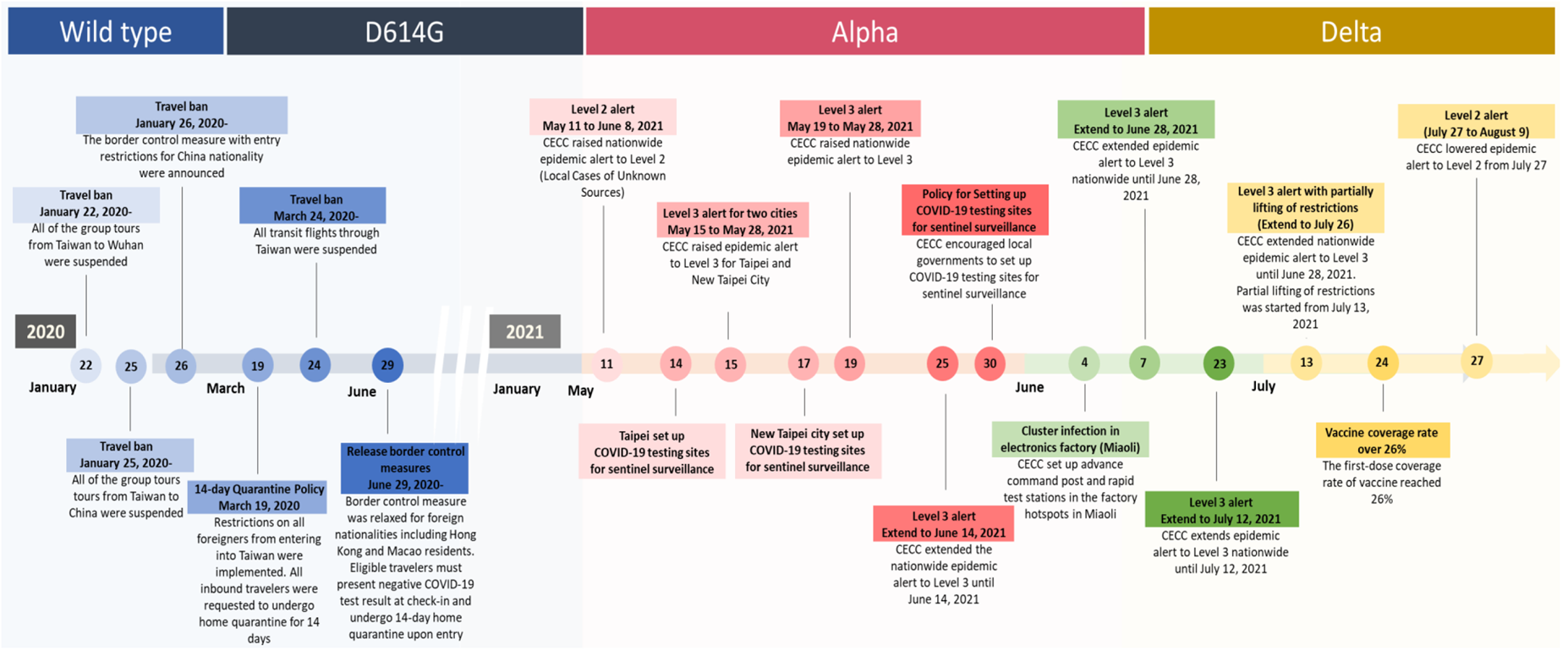
Timelines of SARS-CoV-2 VOC outbreak and the implementation of key containment measures in Taiwan.

**Supplementary table 1.**
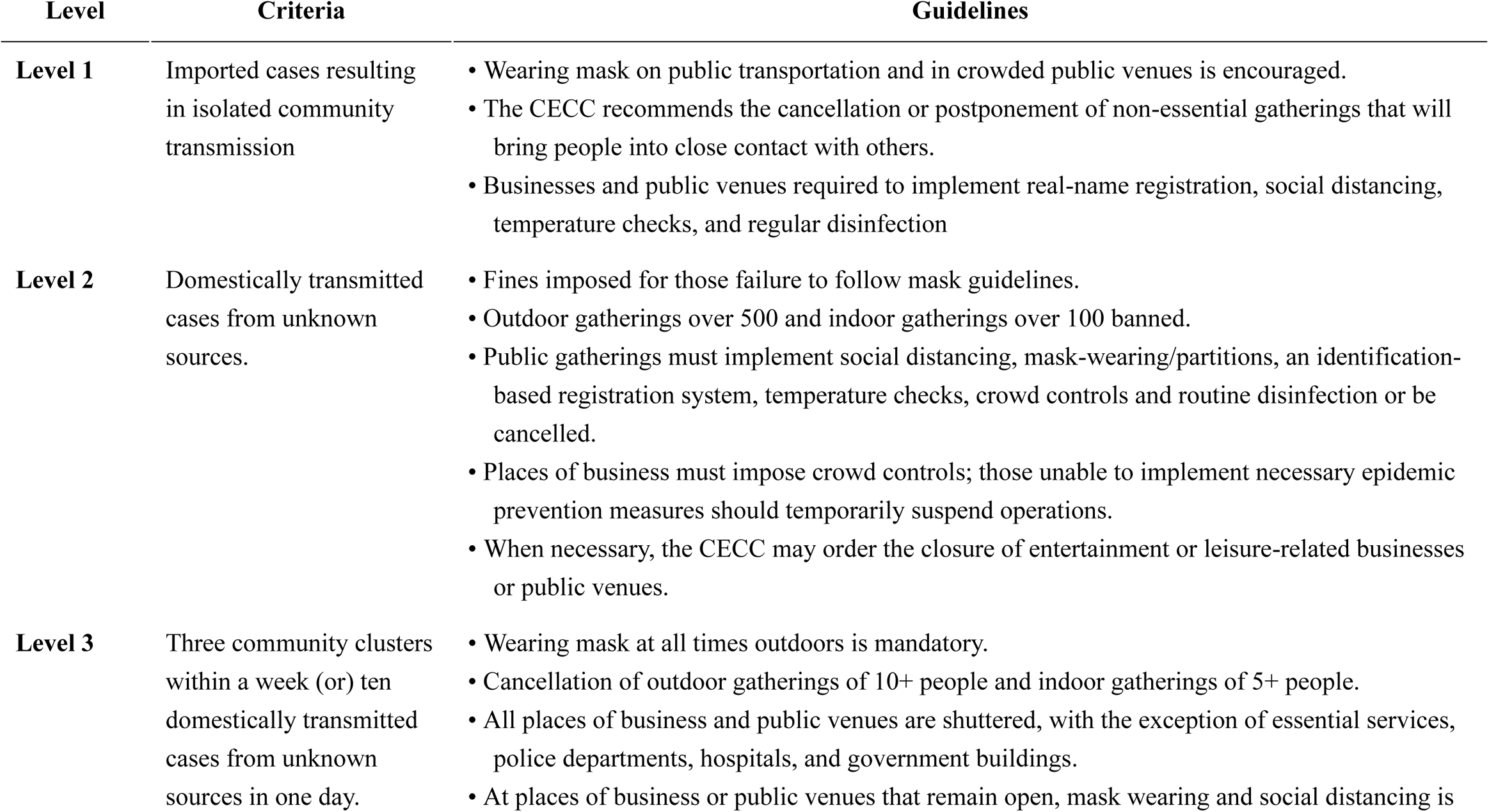
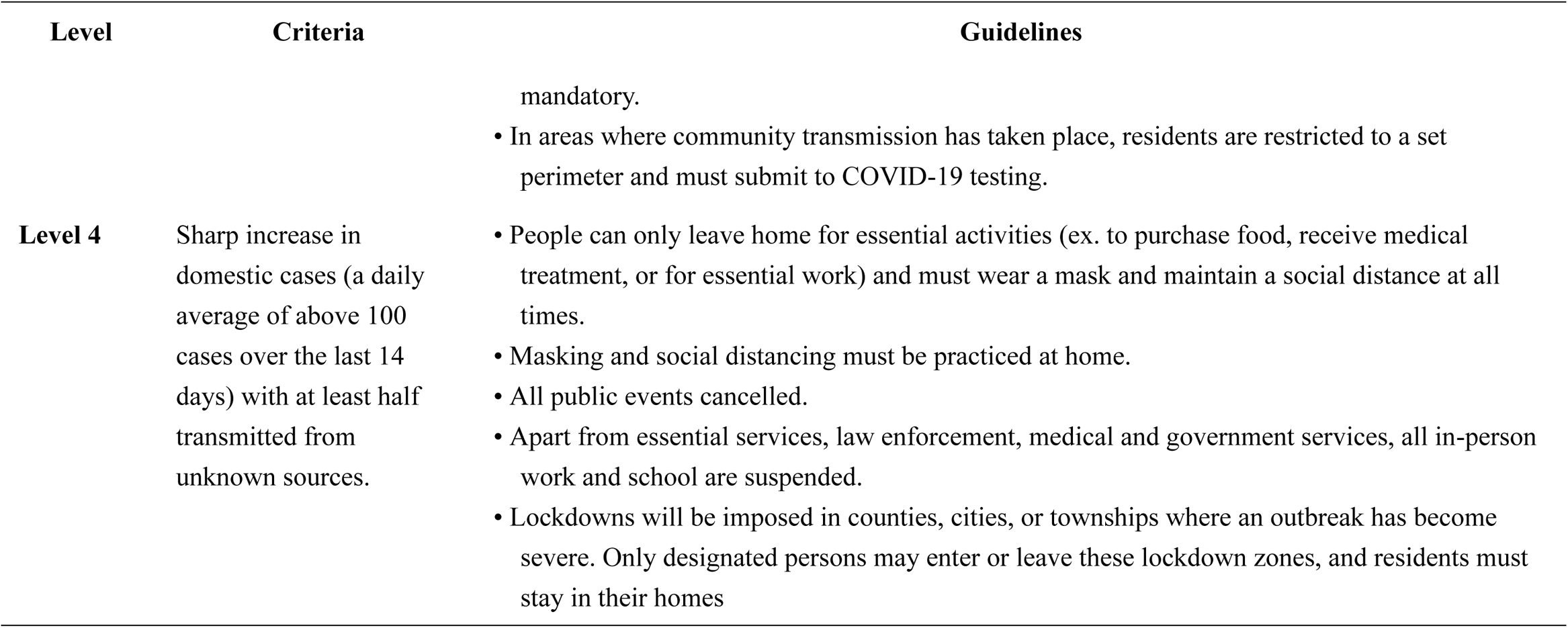
Criteria and guidelines for the four covid-19 alert levels in Taiwan

### B. CAO in Taiwan

**Supplementary figure 2.**
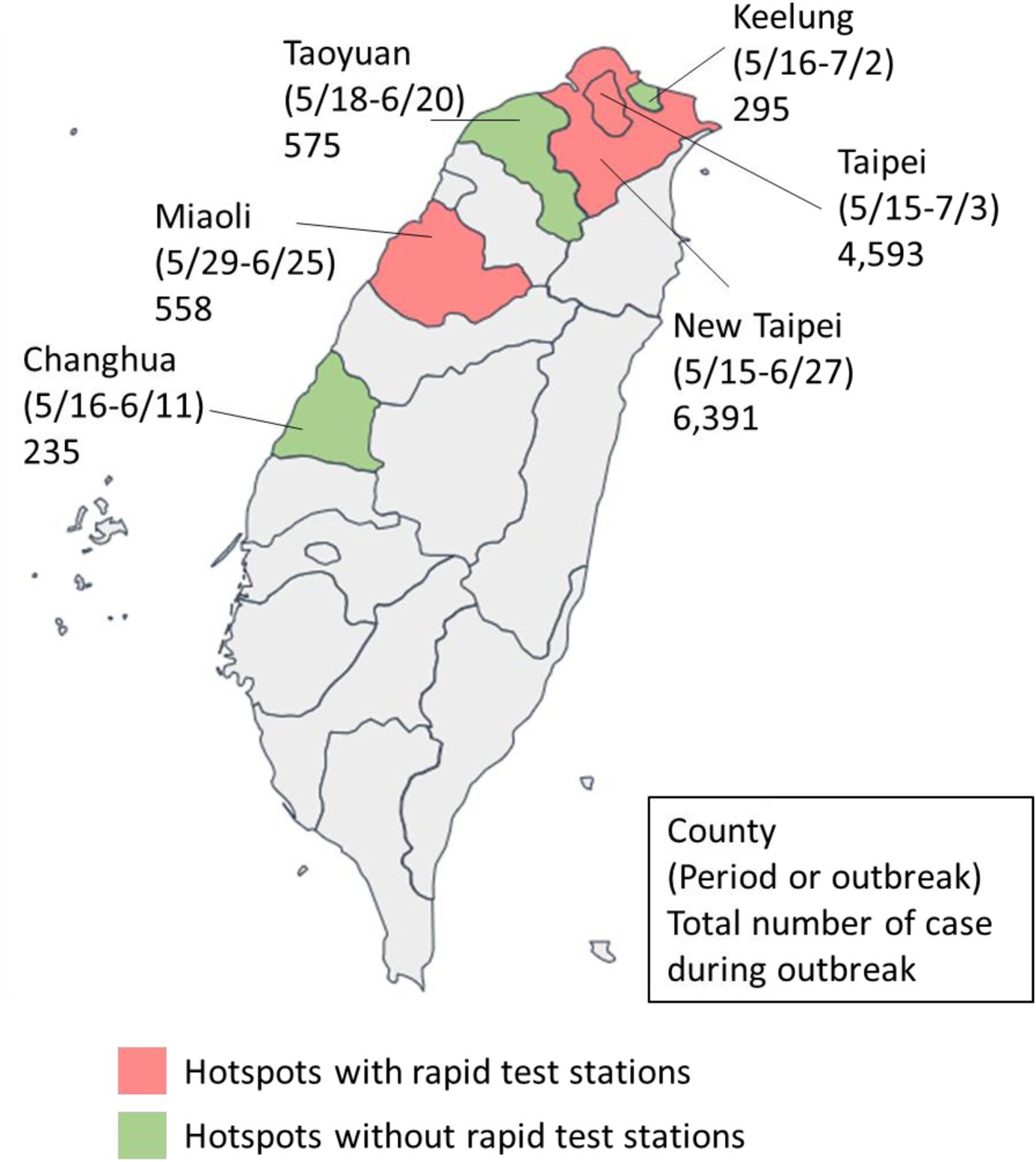
Cities and counties with hotspots in Taiwan

**Supplementary figure 3.**
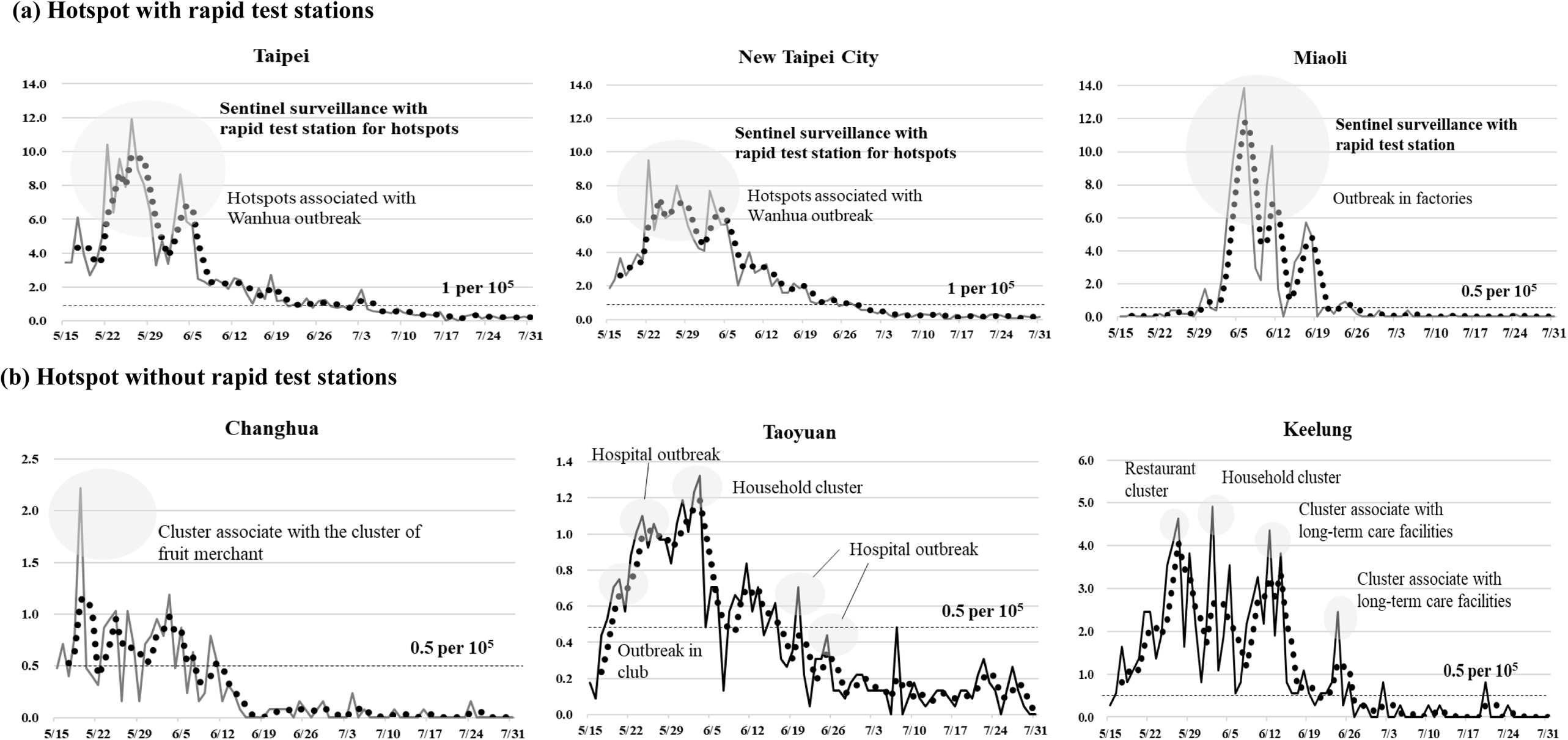
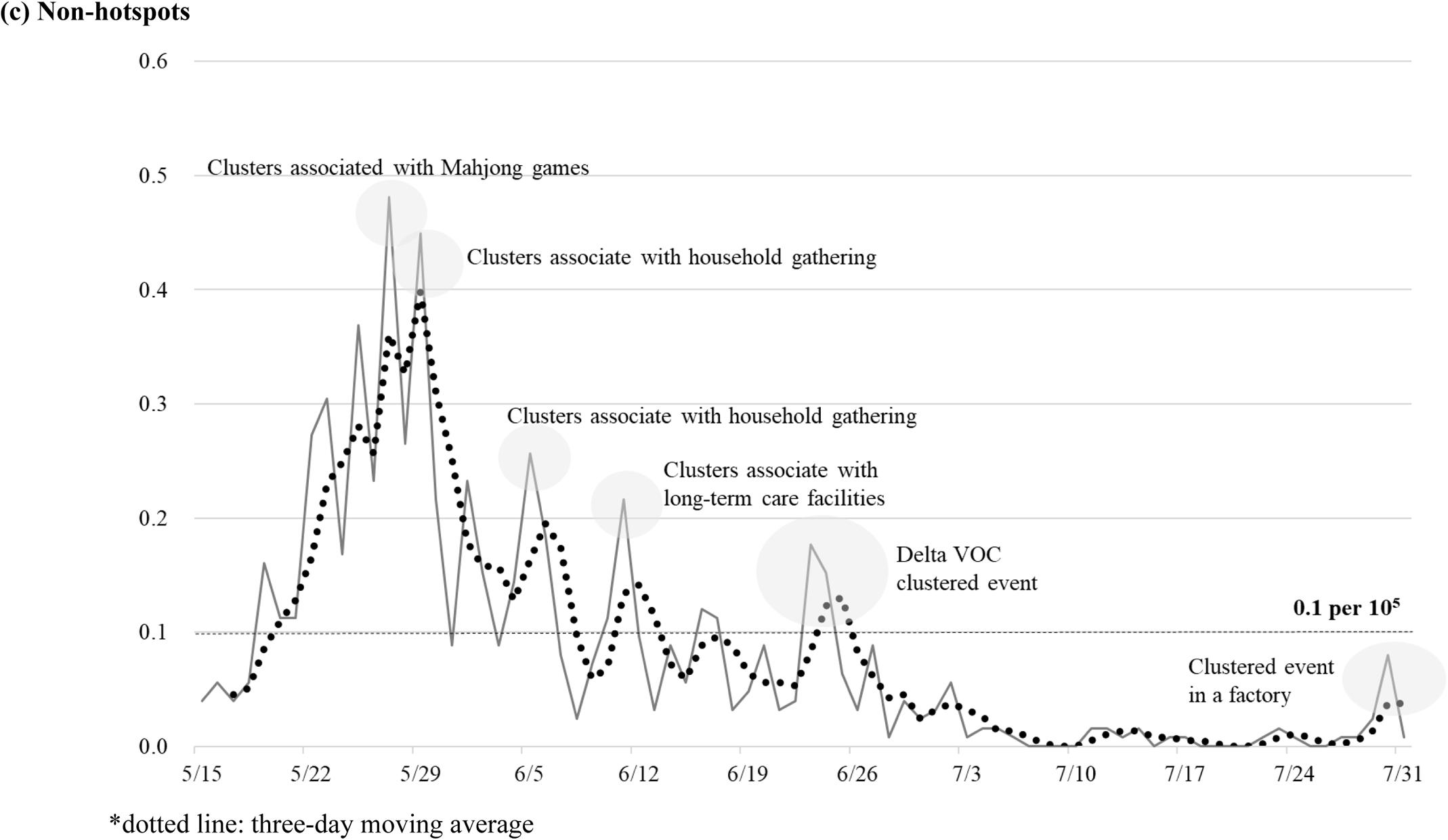
Incidence trend by three categories, hotspots and clustered infections in Taiwan*

**Supplementary table 2.** Estimated results on the time-varying transmission coefficient, reproductive number, and the effectiveness of NPIs and testing by each epoch of community-acquired outbreak in Taiwan in 2021.

| **Date** | **Number of**<br>**reported**<br>**covid-19 cases** | **$\beta_t$** | **$R_t$** | **Effectiveness**<br>**of NPIs and**<br>**testing (%)** |
| --- | --- | --- | --- | --- |
| **16-18 May** | 1275 | 0.58 | 4.0 | 30 |
| **19-22 May** | 1906 | 0.45 | 3.1 | 46 |
| **23-26 May** | 2024 | 0.28 | 2.0 | 66 |
| **27-29 May** | 1359 | 0.14 | 1.0 | 83 |
| **30 May-1 June** | 1152 | 0.17 | 1.2 | 79 |
| **2-4 June** | 1249 | 0.13 | 0.9 | 84 |
| **5-7 June** | 1022 | 0.07 | 0.5 | 92 |
| **8-10 June** | 756 | 0.09 | 0.6 | 89 |
| **11-13 June** | 710 | 0.07 | 0.5 | 91 |
| **14-16 June** | 484 | 0.06 | 0.4 | 93 |
| **17-19 June** | 490 | 0.07 | 0.5 | 92 |
| **20-23 June** | 260 | 0.03 | 0.2 | 96 |
| **24-26 June** | 283 | 0.07 | 0.5 | 91 |
| **27-29 June** | 202 | 0.05 | 0.3 | 94 |
| **30 June-2 July** | 159 | 0.05 | 0.3 | 94 |
| **3-5 July** | 170 | 0.06 | 0.4 | 93 |
| **6-8 July** | 84 | 0.03 | 0.2 | 96 |
| **9-11 July** | 91 | 0.05 | 0.3 | 94 |
| **12-14 July** | 68 | 0.05 | 0.3 | 94 |
| **15-17 July** | 51 | 0.04 | 0.3 | 95 |
| **18-20 July** | 48 | 0.06 | 0.4 | 93 |
| **21-23 July** | 69 | 0.12 | 0.9 | 85 |
| **24-26 July** | 46 | 0.07 | 0.5 | 91 |
| **27-30 July** | 50 | 0.10 | 0.7 | 88 |

\*The transmission coefficient without NPIs and testing ( $\beta_0$ ) was estimated as 0.82 ( $R_0=5.775$ ) based on the data during the initial outbreak (23 April-15 May).

**Supplementary figure 4.**
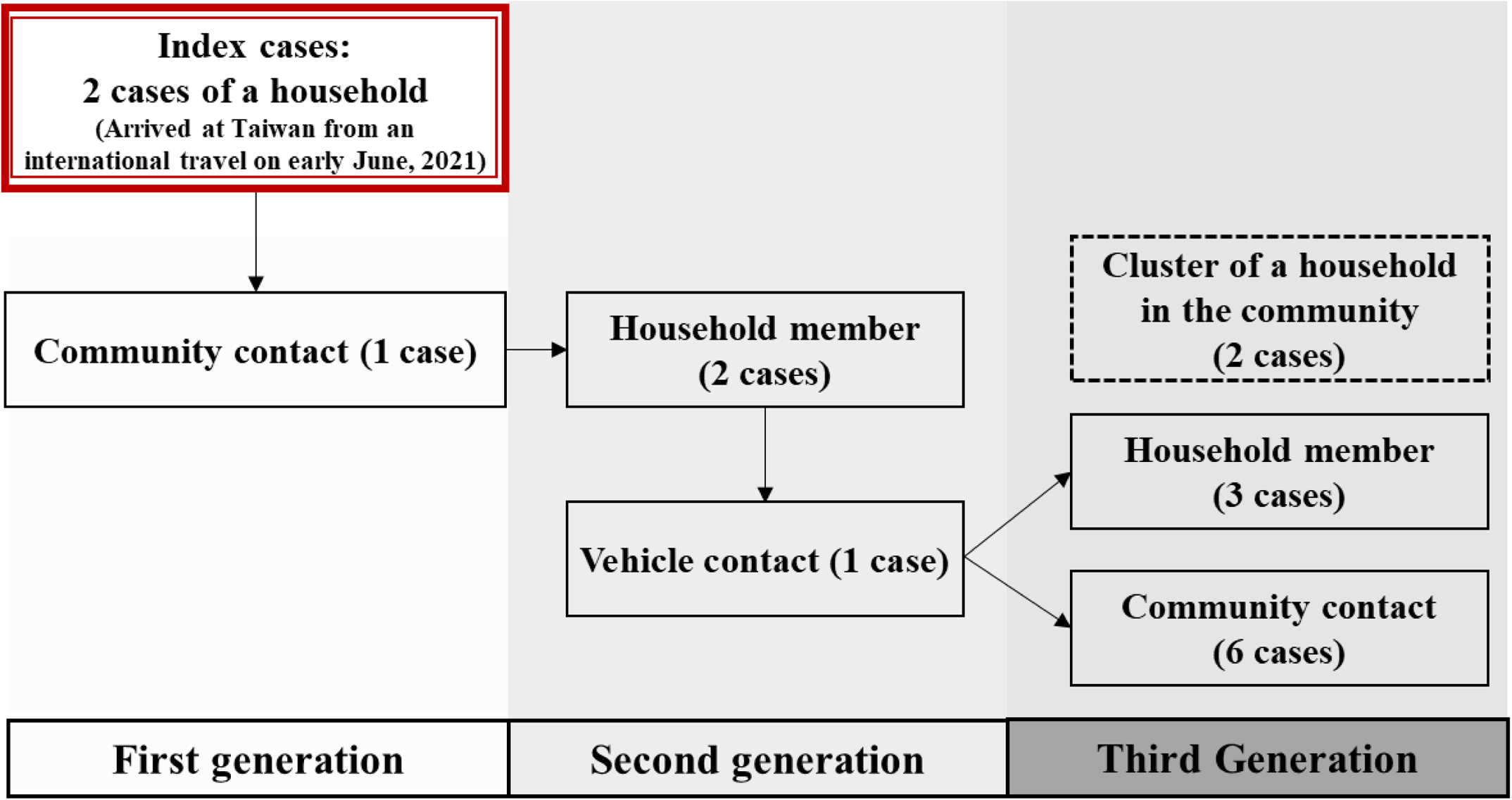
Delta clustered event in Pingtung, Taiwan

**Supplementary figure 5.**
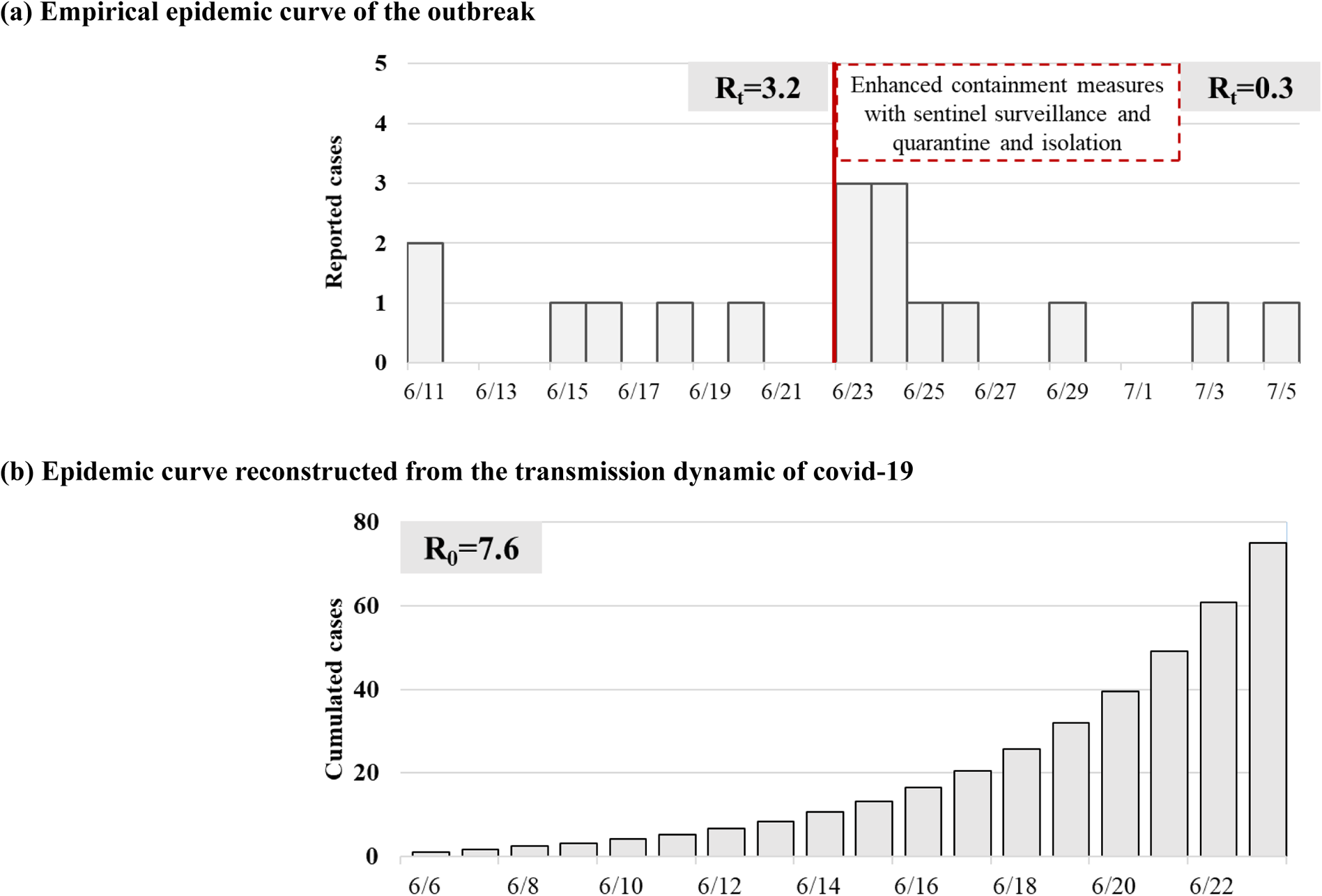
Epidemic curve of the clustered event caused by Delta VOC in Pingtung, Taiwan

### C. External validation of epidemic surveillance model for the imported-domestic transmission without CAO by using data on covid-19 outbreak in New Zealand

Supplementary figure 6 depicts the epidemic curve based on the empirical data from New Zealand. There was only one main outbreak of domestic cases between 29 March and 5 April 2020, which was followed by the peak of imported cases between 22 and 29 March 2020.

Supplementary table 3 shows the association between domestic cases lagging one-week behind imported cases in New Zealand (DIC=1687.9), which is smaller than the model with concurrent-week imported cases (DIC=2173.2) and those lagging two-week behind imported cases (DIC=2395.4) (data not shown but available upon requests). It shows that an increase in one imported case would increase the risk of domestic cases by 9.38% (95% CI 8.88% to 9.86%).

Although the scale of case number in Y axis in supplementary figure 7 for New Zealand is remarkably higher than that of fig 2 for Taiwan, patterns of both Taiwanese and New Zealand curves are similar. One exception is that there was one main outbreak of domestic cases (29 March and 5 April 2020) followed by the peak of imported cases (22-29 March 2020) in New Zealand. Had relevant containment and mitigation measures been taken, as predicted by our proposed surveillance model in the week prior to the cluster infection, such a large-scale community-acquired outbreak might have been averted. However, it can be clearly seen that the subsequent cluster infection between 11 April and 14 April 2020 did not lead to large community-acquired infections in New Zealand, as similarly noted in Taiwan due to the adoption of containment and mitigation measures before cluster infection.

As far as the results of external prediction are concerned, there were three weeks between 9 August and 29 August 2020 in which the observed numbers of domestic cases were beyond the upper limit of surveillance level. The community outbreak in Auckland contributed the majority of this episode. The immediate response to take the measure of lockdown in Auckland successfully curbed the further spread of covid-19.

**Supplementary table 3.**
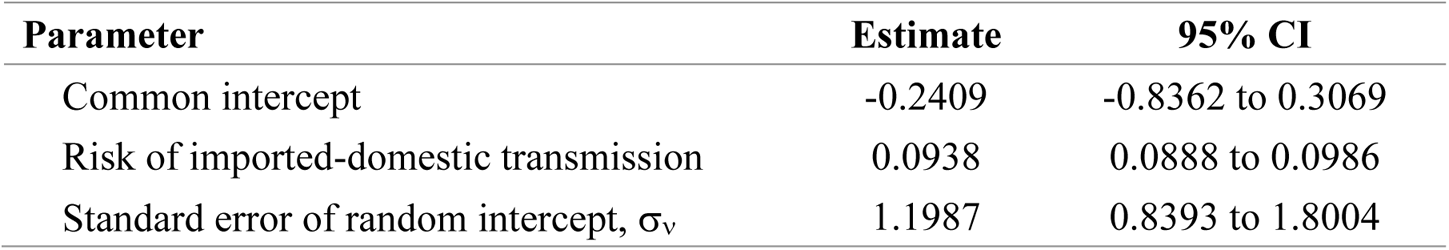
Estimated results for the risk of imported-domestic transmission of covid-19 in New Zealand with the consideration of heterogeneity across counties using Poisson model

| **Parameter** | **Estimate** | **95% CI** |
| --- | --- | --- |
| Common intercept | -0.2409 | -0.8362 to 0.3069 |
| Risk of imported-domestic transmission | 0.0938 | 0.0888 to 0.0986 |
| Standard error of random intercept, $\sigma_v$ | 1.1987 | 0.8393 to 1.8004 |

**Supplementary fig 6.**
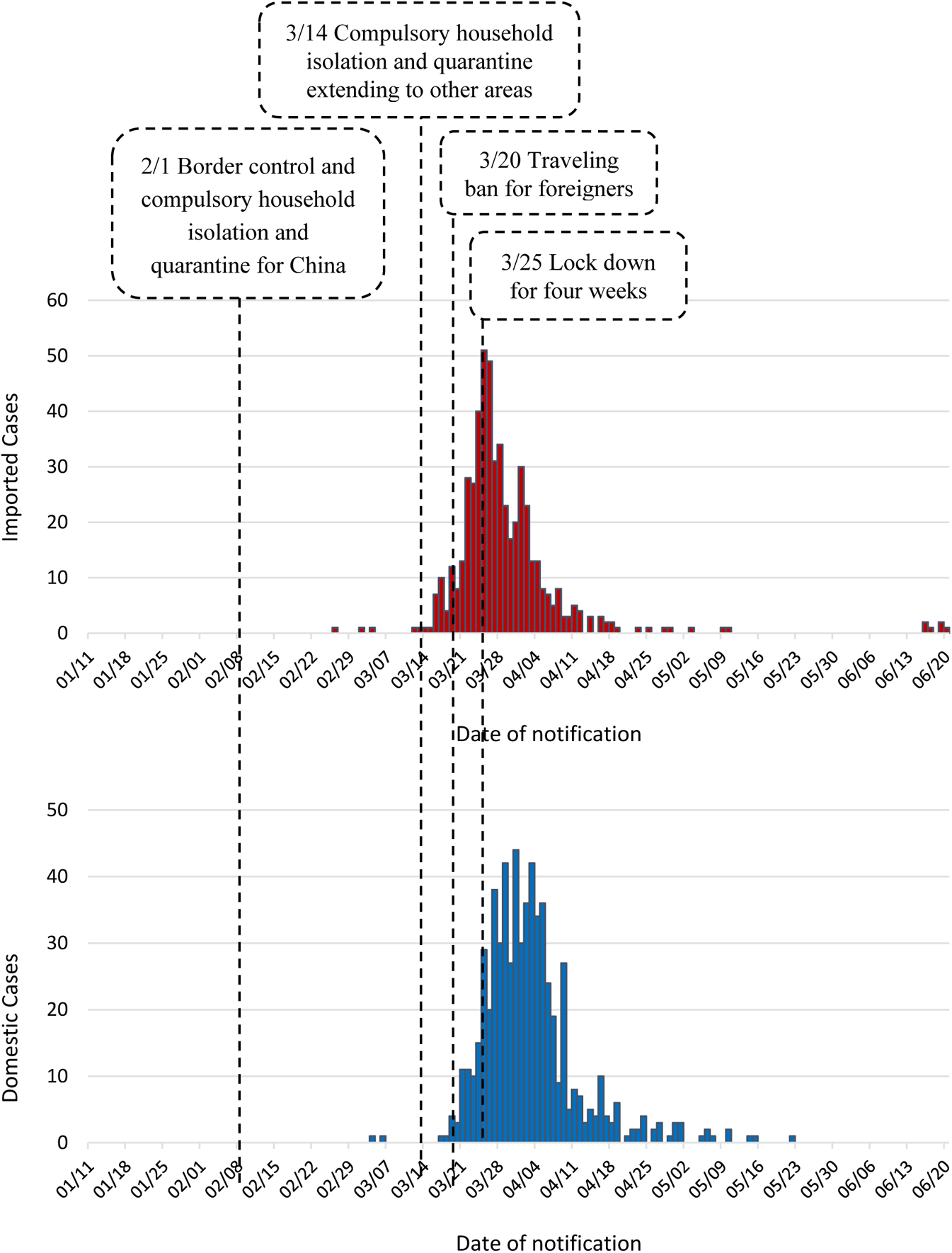
Epidemic curve of covid-19 outbreak in New Zealand by types of cases

**Supplementary fig 7.**
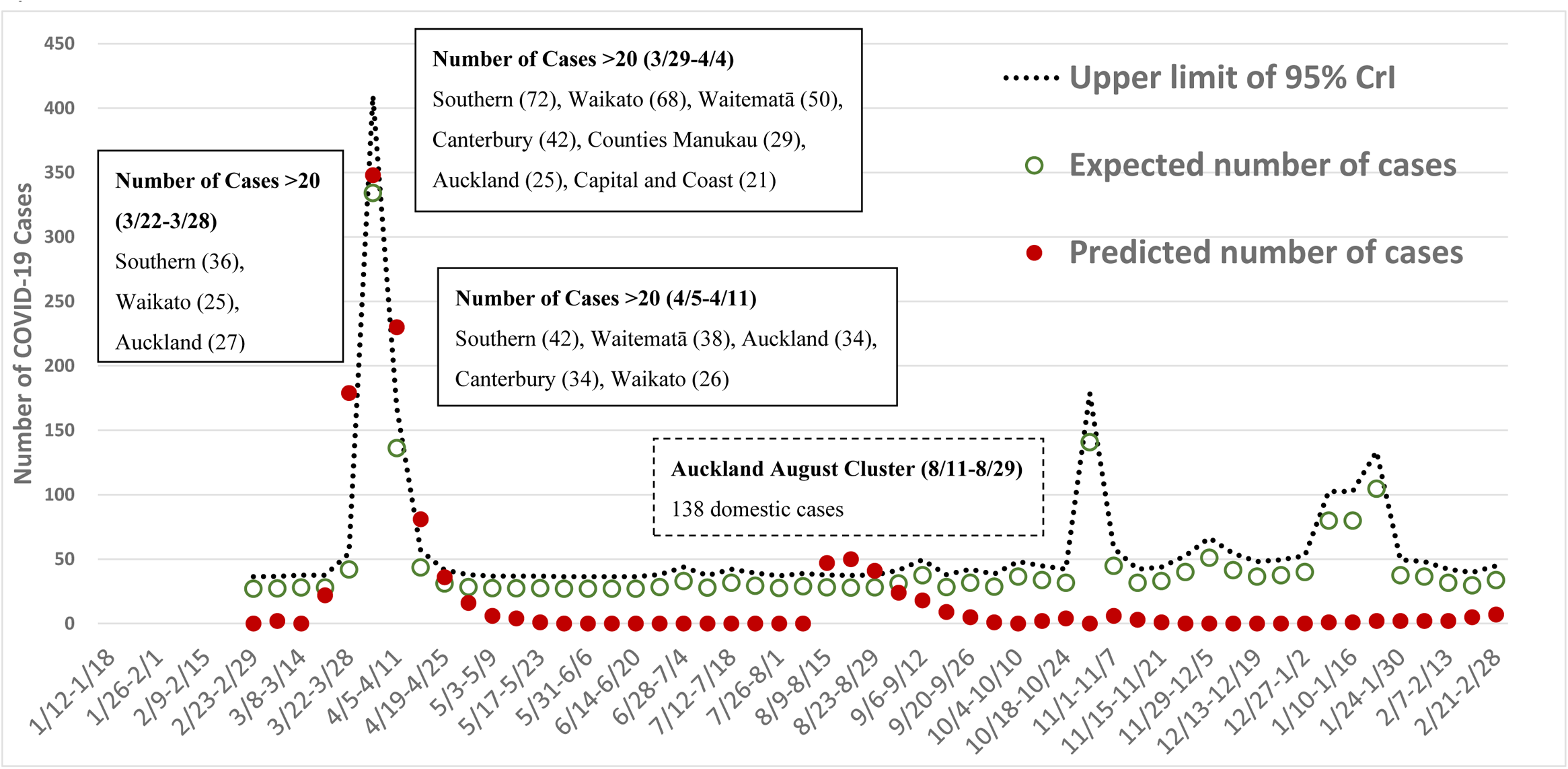
Number of observed (blue dot) and expected (green circle) domestic cases with upper limit of 95% CrI (dotted line) by week

